# Development of Computational Pipeline for Right Ventricular Hemodynamic Single-Beat Analysis

**DOI:** 10.1101/2025.06.01.25328762

**Authors:** Gyeongtae S. Moon, Paul J. Scheel, Maggie Montovano, Milan Kaushik, Cole Buchanan, Samuel H. Friedman, Rebecca Vanderpool, M. Imran Aslam, Ryan J. Tedford, Paul M. Hassoun, Vivek P. Jani, Steven Hsu

## Abstract

**Background:** Load-independent indices of right ventricular (RV) dysfunction aid in the prognosis of patients with pulmonary hypertension (PH), but their acquisition remains difficult. This study aimed to develop a novel computer vision artificial intelligence-based pipeline that can estimate load-independent RV functional indices using screenshots of the RV pressure-time waveform from a standard clinical right heart catheterization (RHC).

**Methods:** Prospectively collected clinical data and research-grade pressure-volume-time data were collected from 76 patients from three centers. Patients were referred for RHC for known or suspected PH. Thirty-nine patients from one center were used for internal development, with external validation performed on the remaining 37 patients from two independent centers. A MATLAB-based computational pipeline was developed to predict RV pressure-volume (P-V) loop and extract load-independent RV indices using image processing and single-beat analysis. Agreement with gold-standard single-beat analysis was assessed via Bland-Altman analysis, concordance correlation coefficient (CCC), and Pearson correlation coefficient. Kaplan-Meier survival analysis, Cox regression analysis, and K-means clustering were performed to evaluate prognostic value.

**Results:** The average age of the derivation cohort was 57±12 years. Strong concordance was observed between the novel and gold-standard methods for end-systolic elastance (Ees, R=0.96, CCC=0.58), effective arterial elastance (Ea, R=0.97, CCC=0.88), end-diastolic elastance (Eed, R=0.87, CCC=0.47), and Ees/Ea ratio (R=0.93, CCC=0.71). Agreement was validated with the external cohort. Prognostic analyses showed that pipeline-derived Ea (HR: 2.09 [1.04, 4.20]) and Ees/Ea (HR: 0.27 [0.08, 0.87]) were significant predictors of clinical outcomes. Cluster analysis identified two RV sub-phenotypes with distinct hemodynamic features, with the group exhibiting higher Ea (1.05±0.27 vs. 0.41±0.19 mmHg/mL, p<0.0001) and lower Ees/Ea (0.37±0.15 vs. 0.76±0.40 mmHg/mL, p=0.0002) demonstrating worse outcomes.

**Conclusion:** We have developed a novel computational pipeline tool that digitizes and generates single-beat estimates of RV-pulmonary arterial coupling from an image of the RV pressure waveform. Its output correlates with single-beat methods and predicts clinical outcomes.

## INTRODUCTION

Right ventricular failure (RVF) is a highly morbid complication of pulmonary hypertension (PH), be it from left heart failure or pre-capillary pulmonary hypertension.(1–5) Our ability to identify, longitudinally assess, and therapeutically target RVF remains limited. Thus, PH and HF patients who develop RVF suffer poorer clinical outcomes.(6–8) Despite its importance, our current clinical assessment of the RV relies on imprecise, load-dependent indices of RV function derived from echocardiography and invasive hemodynamic measurements that only identify RVF at its manifest stages.(9–11) Our inability to separate our assessment of intrinsic RV function from its loading conditions makes it difficult to delineate RV compensation from occult RV dysfunction, which limits earlier identification and intervention.(12–14)

The gold-standard method to assess RV function requires a specialized conductance catheter that can acquire simultaneous RV pressure-volume (P-V) data. This facilitates the measurement of load-independent indices of RV-pulmonary arterial function. These measures include RV end-systolic elastance (Ees), a measure of contractility; effective arterial elastance (Ea), which captures the total afterload imposed upon the RV; and their ratio (Ees/Ea), which encapsulates the mechanical efficiency of the RV-pulmonary arterial unit.(2,15) However, these measurements are not practical to obtain in the clinical arena, as they are invasive, technically challenging, and costly. One accessible alternative is the single-beat method for calculating Ees and Ea, which requires only the RV pressure-time tracing.(16) This method estimates a theoretical peak isovolumic pressure (P_max_), which, along with cardiac output measurement and standard numerical manipulation of the pressure waveform(17), allows for the estimation of our two key elastance values. However, this, too, requires specialized equipment to acquire the raw analog data, thus also limiting it to research settings. A link between these advanced methods and our standard clinical practice remains elusive.

In this study, we sought to bridge this divide by leveraging the standard reports of right heart catheterization (RHC) data from the Electronic Health Record to generate clinically relevant RV pressure-volume loop data. To this end, we use computer vision artificial intelligence to develop a new semi-automated pipeline for single beat estimation of load-independent RV contractile indices that requires only the screenshot of an RV waveform from a standard RHC report along with an ancillary measurement of cardiac output (**Central Illustration).** We apply our pipeline to data obtained from three academic clinical centers and compare it against a traditional single beat analysis. We then assess the prognostic value of the newly derived RV pressure-volume indices beyond the standard echocardiographic and hemodynamic assessment of RV function.

## METHODS

### Study Population

The derivation cohort of this study is from a prospectively assessed cohort of 84 patients referred for exercise right heart catheterization for known or suspected PH performed at the Johns Hopkins Hospital between 2013 and 2024.(18) The study protocol was approved by the Johns Hopkins Institutional Review Board, and all patients gave written informed consent. All patients, regardless of PH status, were included so long as they had complete rest hemodynamic measurements, concomitant cardiac MRI (CMR), P-V measurements of the RV at rest, and a digital image of RV pressure tracings. Of these subjects, 39 met the inclusion criteria. Two blinded cardiac radiologists (S.L.Z. and B.A.V.) analyzed the CMR findings. These included patients with connective tissue disease-associated versus idiopathic pulmonary arterial hypertension (IPAH). PH was identified as a mean PA pressure (mPAP) > 20 mmHg. Additional criteria for Group 1 PAH included pulmonary vascular resistance > 2 Wood units and pulmonary artery wedge pressure (PAWP) ≤ 15 mmHg(19). Group 2 PH was classified as mPAP > 20 mmHg and PAWP *>* 15 mmHg.(19) PAH was adjudicated as by specialists (C.E.S., T.M.K., R.I.D., S.C.M., P.M.H.), as described previously.(20,21) Detailed clinical and laboratory characteristics were obtained at the time of enrollment.

External validation datasets included subjects with complete rest hemodynamic measurement, analog pressure waveform data, and a digital image of the RV pressure waveform. Data were obtained from two academic centers: 17 subjects referred for right heart catheterization due to known or suspected pulmonary hypertension or unexplained dyspnea from the Medical University of South Carolina (R.J.T.) and 20 subjects with HF and reduced ejection fraction (EF) with cardiac catheterization based on suspicion of Group 2 PH from Duke University (M.I.A.).

### Right Heart Catheterization and Conductance Catheter Measurements

Patients underwent standard RHC procedure, simultaneous RV P-V loop measurements, and same-day CMR using a previously described protocol.(22) Following the RHC procedure, a P-V conductance catheter (CA-41103-PN, CD Leycom, Netherlands) was placed in in the RV under fluoroscopic guidance. Steady-state data were obtained at end-expiration at baseline. Analog pressure waveforms were obtained and sampled at 250 Hz for traditional single beat analysis. All collected raw data was sent to the laboratory for further processing and analysis.

### Development of Computational Pipeline for Single Beat Analysis

A MATLAB (The MathWorks, Inc, US) application, available online here, was developed to allow for a semi-automatic extraction of single beat P-V loop parameters with an image of RV pressure tracing.(17,23) With the application, waveform reports were digitized using various image processing and computer vision artificial intelligence techniques (including notch filtering, canny edge detection, morphological operations, and median filtering). Then, they were subsequently averaged for single-beat analysis. Maximal pressure (P_max_) was calculated using a piece-wise sinusoidal fit to dP/dt_max_ and dP/dt_min_ with a set of nonlinear equations (see Appendix) to allow for more consistent acquisition of P_max_. Stroke volume (SV) is derived from Fick or thermodilution cardiac output (CO) with a given heart rate. End-systolic (ESP) and end-diastolic pressures (EDP) were estimated using the square of the second derivative of the waveform.(17) The end-diastolic pressure-volume relationship (EDPVR) was calculated by fitting an exponential function through (0, 0), (ESV, 1), and (EDV, EDP). Ejection was estimated using a cubic spline between (ESV, ESP) and (EDV, Pressure[dP/dt_max_]). RV functional parameters Ees (end-systolic elastance), Ea (effective arterial elastance), Eed (End diastolic elastance), and RVEF (right ventricle ejection fraction) were obtained as previously described.(3,17) Logistic tau was obtained numerically as previously described.(24)

### Statistical Approach

Results are expressed as mean ± standard deviation unless otherwise stated. Variables between groups were compared using the Mann-Whitney two-sample non-parametric test. The primary endpoint was a combined endpoint of death, lung transplant, or clinical worsening (defined as either a HF hospitalization, worsening New York Heart Association functional classification, 15% decline in 6-minute walk test, or escalation of PAH therapy). These were collected for the derivation cohort. A non-parametric right-censored life table analysis was conducted using Kaplan-Meier estimator to construct survival curves. Hazard ratios for the primary endpoint were estimated using Cox proportional hazards regression models adjusting for relevant confounders (PVR, BMI, age, sex, race, GFR). Data were tested for normality using the Shapiro-Wilk test and log-transformed when needed. Agreement for clinical variables obtained through the computational pipeline vs. gold standard was evaluated using Bland-Altman analysis, concordance correlation coefficient (CCC), and Pearson coefficient (R). Based on variables extracted from single beat analysis, the 39 patients from the derivation cohort were stratified into two groups using the K-Means clustering algorithm to identify subgroups with distinct clinical characteristics that might show prognostic significance (Python, scikit-learn ver. 1.3.0). Statistical significance was defined by a two-sided p-value < 0.05. Statistical analyses were performed using Stata 15.1 (StataCorp LLC, TX) and R Statistical Software (v. 4.3.1, R Core Team).

## RESULTS

### Baseline Patient Characteristics

The study population in the derivation cohort consisted of 39 patients with invasive hemodynamics who were predominantly Caucasian (27/39, 69.2%) and female (36/39, 92.3%), with a mean age of 57±12 years. Eight of 39 (20.5%) were newly diagnosed with PAH after RHC, while 22 (56.4%) had prevalent PAH. 5 (12.8%), 26 (66.7%), and 8 (20.5%) were in WHO Classes 1, 2, and 3, respectively. Among the cohort, 12 (31%) subjects had idiopathic PAH (iPAH). Median NT-proBNP was 222 pg/mL (136-397, 95% CI). On right heart catheterization, the mean right atrial pressure (RAP) was 6 ± 2.6 mmHg, the mean pulmonary artery pressure (mPAP) 31 ± 12 mmHg, pulmonary vascular resistance (PVR) 5 ± 3 WU, and the mean pulmonary artery wedge pressure (PAWP) 10 ± 4 mmHg. CO and CI were 4.8 ± 1.4 L/min, and CI 2.5 ± 0.6 L/min/m^2^, respectively. Additional characteristics are summarized in **Supplemental Table 1** with external validation cohorts.

### Image-Derived Single Beat Analysis

Using computer vision artificial intelligence, we developed a semi-automated pipeline for the acquisition of Ees, Ea, and Eed from an image of an RV waveform easily accessible by chart review. We then tested the output of this approach against that of traditional single beat analysis in the derivation cohort described above(23,25,26). Our computational pipeline includes an interactive graphical user interface that allows the users to conduct the analysis intuitively. No prior computational experience is required. We first evaluated whether our approach yielded results similar to traditional single beat analysis. All parameters indeed showed strong agreement with parameters obtained using traditional single beat analysis (**Table 1**). Of note, we observed strong agreement for Ees (R=0.96, CCC = 0.58), Ea (R=0.97, CCC=0.88), and their ratio (R=0.93, CCC=0.71). Recently, it has been shown that diastolic elastance (Eed) is significantly associated with cardiac index reserve in Group 1 PAH patients(3). We, therefore, interrogated whether our approach could obtain Eed and indeed also found a moderate agreement versus traditional single beat analysis (R=0.87, CCC=0.47).

**Table 1.**
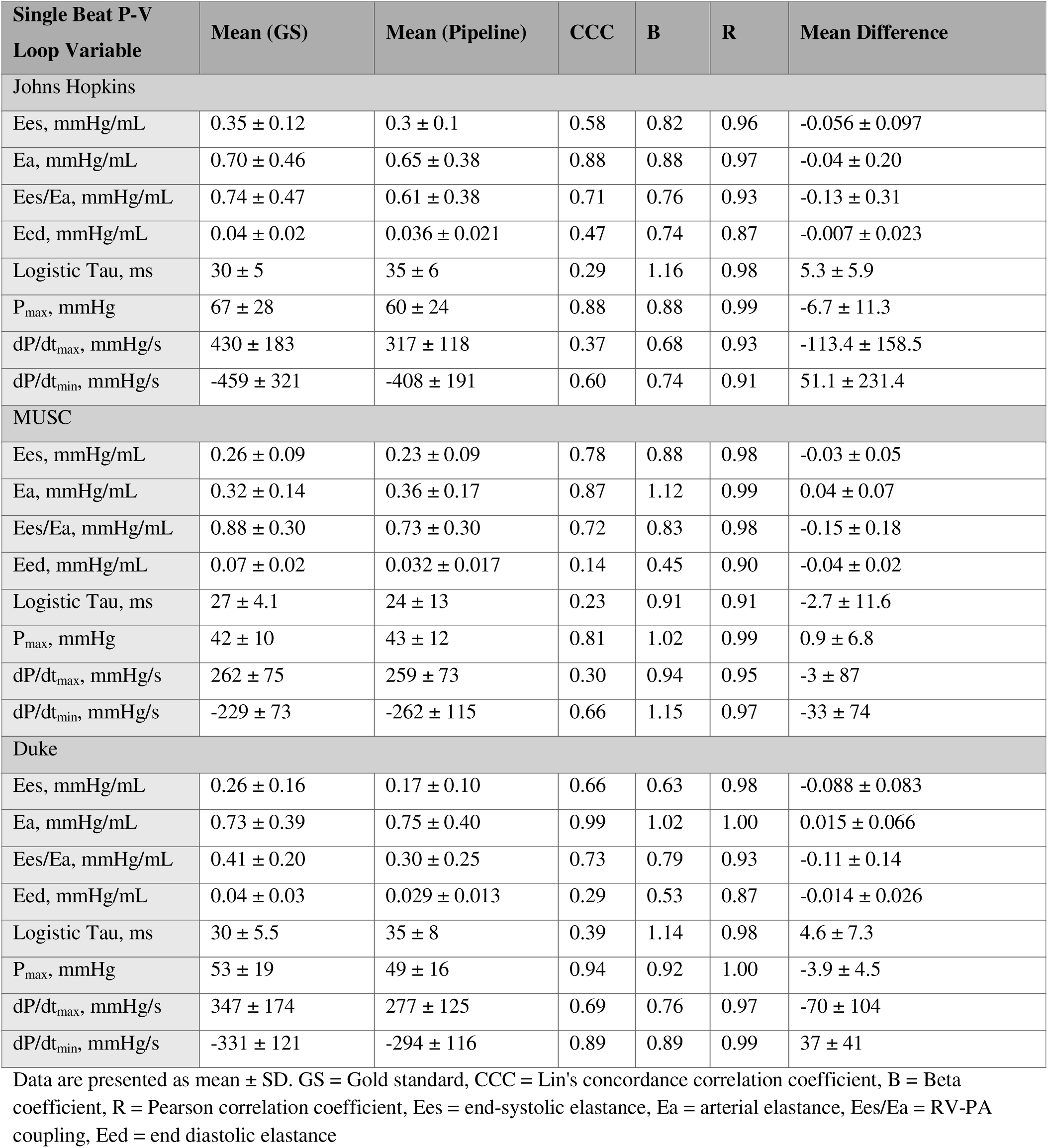
Goodness of Fit Metrics between Gold Standard Single Beat P-V Loop Variables and Pipeline Results in 3 Centers.

| Single Beat P-V Loop Variable | Mean (GS) | Mean (Pipeline) | CCC | B | R | Mean Difference |
| --- | --- | --- | --- | --- | --- | --- |
| Johns Hopkins |  |  |  |  |  |  |
| Ees, mmHg/mL | $0.35 \pm 0.12$ | $0.3 \pm 0.1$ | 0.58 | 0.82 | 0.96 | $-0.056 \pm 0.097$ |
| Ea, mmHg/mL | $0.70 \pm 0.46$ | $0.65 \pm 0.38$ | 0.88 | 0.88 | 0.97 | $-0.04 \pm 0.20$ |
| Ees/Ea, mmHg/mL | $0.74 \pm 0.47$ | $0.61 \pm 0.38$ | 0.71 | 0.76 | 0.93 | $-0.13 \pm 0.31$ |
| Eed, mmHg/mL | $0.04 \pm 0.02$ | $0.036 \pm 0.021$ | 0.47 | 0.74 | 0.87 | $-0.007 \pm 0.023$ |
| Logistic Tau, ms | $30 \pm 5$ | $35 \pm 6$ | 0.29 | 1.16 | 0.98 | $5.3 \pm 5.9$ |
| P<sub>max</sub>, mmHg | $67 \pm 28$ | $60 \pm 24$ | 0.88 | 0.88 | 0.99 | $-6.7 \pm 11.3$ |
| dP/dt<sub>max</sub>, mmHg/s | $430 \pm 183$ | $317 \pm 118$ | 0.37 | 0.68 | 0.93 | $-113.4 \pm 158.5$ |
| dP/dt<sub>min</sub>, mmHg/s | $-459 \pm 321$ | $-408 \pm 191$ | 0.60 | 0.74 | 0.91 | $51.1 \pm 231.4$ |
| MUSC |  |  |  |  |  |  |
| Ees, mmHg/mL | $0.26 \pm 0.09$ | $0.23 \pm 0.09$ | 0.78 | 0.88 | 0.98 | $-0.03 \pm 0.05$ |
| Ea, mmHg/mL | $0.32 \pm 0.14$ | $0.36 \pm 0.17$ | 0.87 | 1.12 | 0.99 | $0.04 \pm 0.07$ |
| Ees/Ea, mmHg/mL | $0.88 \pm 0.30$ | $0.73 \pm 0.30$ | 0.72 | 0.83 | 0.98 | $-0.15 \pm 0.18$ |
| Eed, mmHg/mL | $0.07 \pm 0.02$ | $0.032 \pm 0.017$ | 0.14 | 0.45 | 0.90 | $-0.04 \pm 0.02$ |
| Logistic Tau, ms | $27 \pm 4.1$ | $24 \pm 13$ | 0.23 | 0.91 | 0.91 | $-2.7 \pm 11.6$ |
| P<sub>max</sub>, mmHg | $42 \pm 10$ | $43 \pm 12$ | 0.81 | 1.02 | 0.99 | $0.9 \pm 6.8$ |
| dP/dt<sub>max</sub>, mmHg/s | $262 \pm 75$ | $259 \pm 73$ | 0.30 | 0.94 | 0.95 | $-3 \pm 87$ |
| dP/dt<sub>min</sub>, mmHg/s | $-229 \pm 73$ | $-262 \pm 115$ | 0.66 | 1.15 | 0.97 | $-33 \pm 74$ |
| Duke |  |  |  |  |  |  |
| Ees, mmHg/mL | $0.26 \pm 0.16$ | $0.17 \pm 0.10$ | 0.66 | 0.63 | 0.98 | $-0.088 \pm 0.083$ |
| Ea, mmHg/mL | $0.73 \pm 0.39$ | $0.75 \pm 0.40$ | 0.99 | 1.02 | 1.00 | $0.015 \pm 0.066$ |
| Ees/Ea, mmHg/mL | $0.41 \pm 0.20$ | $0.30 \pm 0.25$ | 0.73 | 0.79 | 0.93 | $-0.11 \pm 0.14$ |
| Eed, mmHg/mL | $0.04 \pm 0.03$ | $0.029 \pm 0.013$ | 0.29 | 0.53 | 0.87 | $-0.014 \pm 0.026$ |
| Logistic Tau, ms | $30 \pm 5.5$ | $35 \pm 8$ | 0.39 | 1.14 | 0.98 | $4.6 \pm 7.3$ |
| P<sub>max</sub>, mmHg | $53 \pm 19$ | $49 \pm 16$ | 0.94 | 0.92 | 1.00 | $-3.9 \pm 4.5$ |
| dP/dt<sub>max</sub>, mmHg/s | $347 \pm 174$ | $277 \pm 125$ | 0.69 | 0.76 | 0.97 | $-70 \pm 104$ |
| dP/dt<sub>min</sub>, mmHg/s | $-331 \pm 121$ | $-294 \pm 116$ | 0.89 | 0.89 | 0.99 | $37 \pm 41$ |

Data are presented as mean ± SD. GS = Gold standard, CCC = Lin's concordance correlation coefficient, B = Beta coefficient, R = Pearson correlation coefficient, Ees = end-systolic elastance, Ea = arterial elastance, Ees/Ea = RV-PA coupling, Eed = end diastolic elastance

We subsequently applied our algorithm to two external cohorts, one group of 17 patients with preserved EF MUSC) and another of 20 patients with Group 2 PH (Duke). In addition to the application of our process to patients with PH related to left-sided heart failure, we also sought to apply our software to the very visually distinct clinical reports derived from the latter institution. In both validation cohorts, our pipeline showed strong agreement in Ees (MUSC: R=0.98, Duke: R=0.98), Ea (MUSC: R =0.99, Duke: R=1.00), and Ees/Ea ratio (MUSC: R=0.98, Duke: R=0.93). Summary results are shown in **Figure 2**. Bland-Altman (**Supplement Figure 1**) was also performed in all three cohorts. For all parameters, minimal bias was observed. The variance was even across the range of values in all three cohorts, except for Ees in the derivation cohort, showing higher variance at higher values.

**Figure 1.**
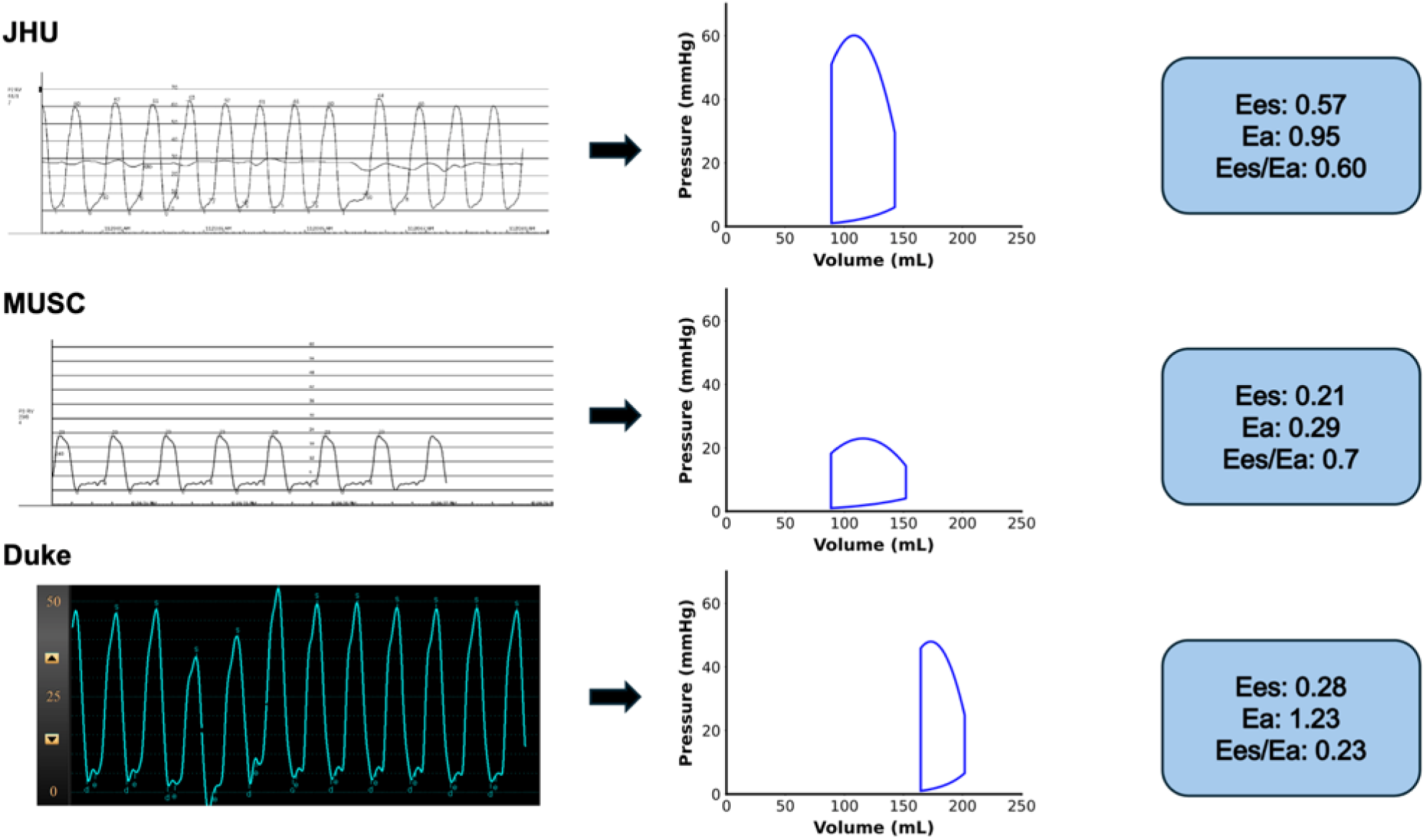
Single-Beat Analysis and P-V Loop Successfully Performed for RV Pressure-Time Tracings from Different Centers. Representative RV pressure-time tracings are shown from different cohorts in different centers (JHH, MUSC, Duke). The pressure-time tracing images were converted into an average waveform, which were then used to estimate the RV P-V loop and single beat parameters.

**Figure 2.**
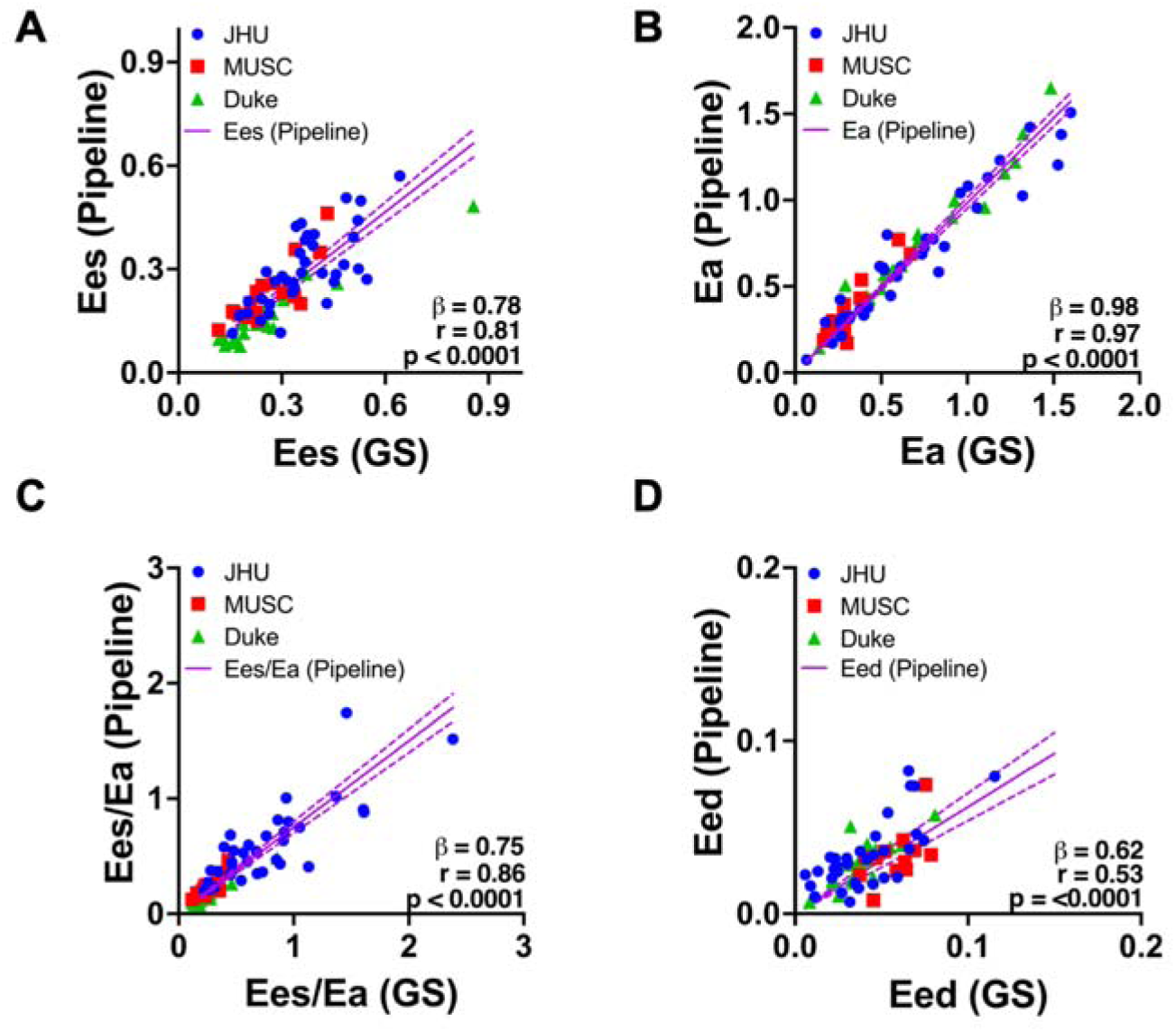
Regression Analysis of Ees, Ea, Ees/Ea, Eed in Different Cohorts between Gold Standard (GS) and Computational Pipeline Outputs. Beta coefficient (β), Pearson r (r), and P values for correlation are shown for each analysis. Linear regression lines are shown as mean with 95% CI. The load-independent indices of RV contractility obtained from the computational pipeline (Ees (**2A**), Ea (**2B**), Ees/Ea (**2C**), Eed (**2D**)) show strong agreement with results obtained from the gold standard method.

### Single Beat-Derived Predictors of Clinical Outcomes

We next performed Kaplan-Meier analysis of the derivation cohort to assess whether single-beat derived indices show prognostic significance (**Table 2**). To test this, we performed Cox regression to identify predictors of our combined endpoint of death, lung transplant, or clinical worsening, as defined in the methods. We find that Ea (HR: 2.09 [1.04, 4.20]), RVESP (HR: 0.33 [0.18, 0.16)], P_max_ (1.90 [HR: 1.08, 3.36]), and dP/dt_min_ (HR: 0.29 [0.14, 0.61]) all significantly associate with our combined endpoint. This is concordant with Cox regression analyses using traditional single beat indices. Also of note, Ees/Ea ratio alone was found to be a significant predictor of HF hospitalization in our cohort (HR: 0.27 [0.08, 0.87]).

**Table 2.**
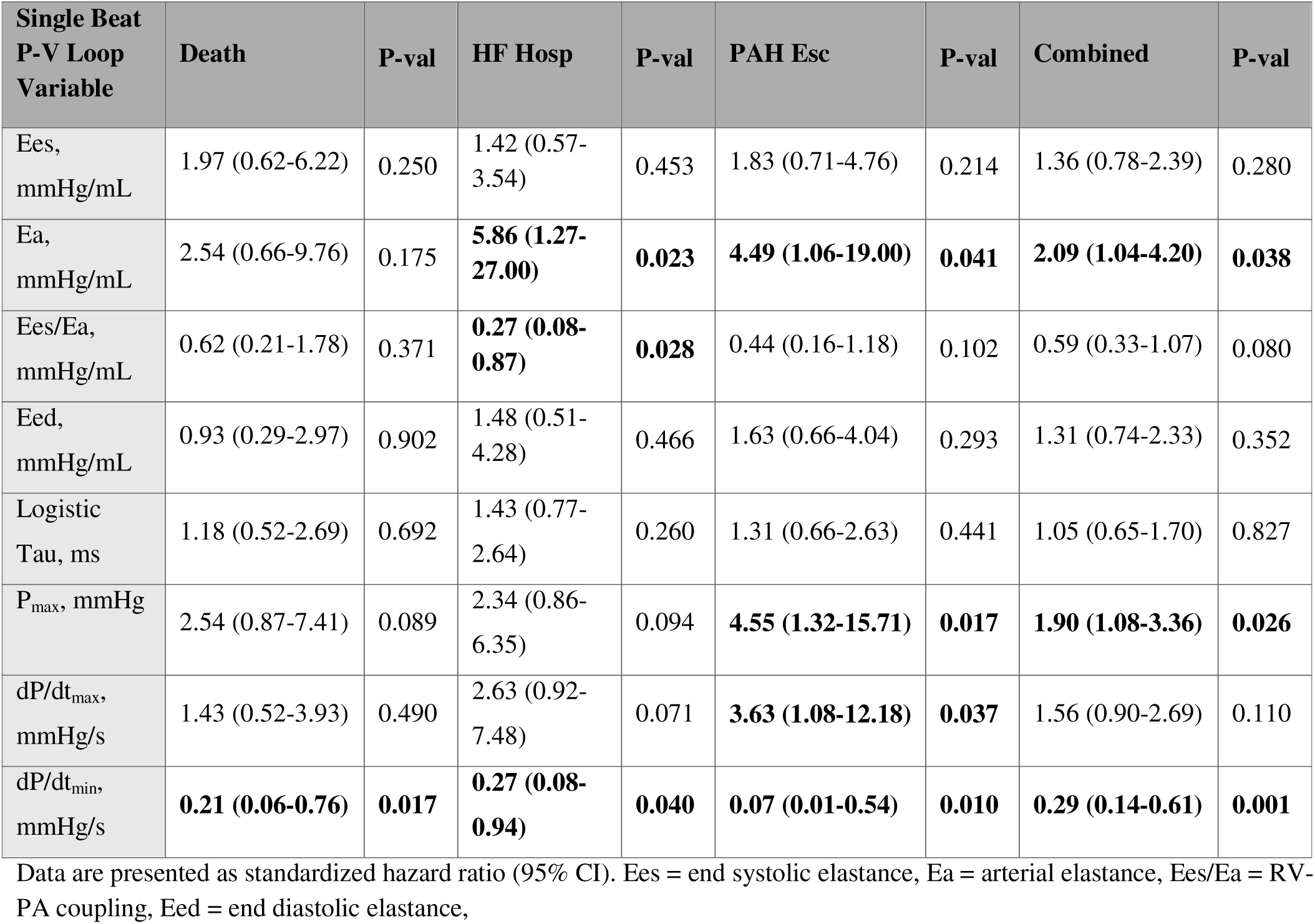
Relationship between Single Beat Parameters from Pipeline and Outcome Variables as Standardized Hazard Ratio (95% CI).

### Cluster Analysis of SB Hemodynamic Phenotypes

We subsequently tested whether sub-phenotypes of RV contractile function, as assessed by single beat analysis, proved clinically significant on their own. Cluster analysis identified two subgroups of RV function, with one group (Group A) exhibiting slightly higher Ees (0.35±0.10 vs. 0.26±0.11 mmHg/mL, p=0.0073) but even higher Ea (1.05±0.27 vs. 0.41±0.19 mmHg/mL p<0.0001), thus leading to significantly lower Ees/Ea (0.37±0.15 vs. 0.76±0.40 mmHg/mL, p = 0.0002) than the other (Group B, **Figure 3A**, **3B**, **Table 3**). We next compared the clinical features of our two groups. In concordance with clinical data, Groups A and B exhibited distinct differences in their clinical hemodynamic profile and structural abnormalities as derived from CMR imaging (**Table 4**). Group A patients show relatively greater mPAP (43.0±7.1 vs. 22.6±6.8, p<0.0001), PVR (8±2 vs. 3±2, p<0.0001), RVEF depression on CMR (43±10 vs. 53±8 %, p=0.002), and RV hypertrophy (RV Mass Index: 36±10.7 vs. 28±9.3, p=0.02), but no change in CI (2.4±0.5 vs. 2.6±0.7 mL/min/m^2^, p=0.48) (**Table 4**). Interestingly, Group A patients were more likely to have prolonged logistic tau (39±6 vs. 33±4, p=0.001). Upon Cox regression analysis of our combined endpoint, we find that patients in the pipeline-derived Group A show an increased hazard ratio (6.047 [1.780, 20.54], versus those in Group B; overall, the groupings could powerfully predict 24-month outcomes (p=0.0001) (**Figure 3C**). Furthermore, we examined whether including single-beat variables in the standard clinical metrics like RHC and CMR imaging variables could improve prognostic assessment. The two resulting groups separated 24_month outcomes significantly (p=0.0298), whereas groups derived from clinical variables alone (RHC and CMR) showed no prognostic discrimination (p=0.4347).

**Figure 3.**
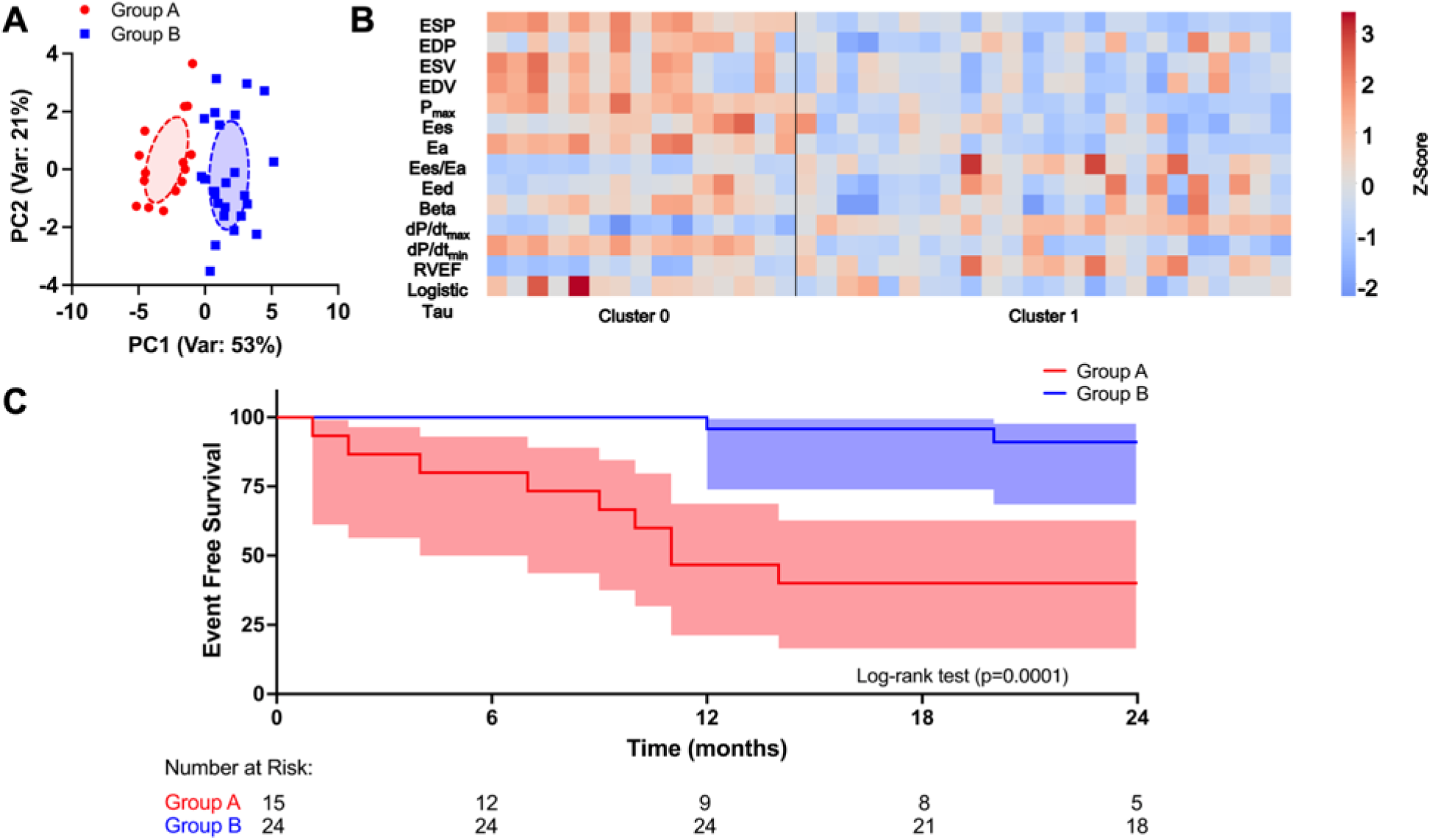
Cluster Analysis with Single Beat Pipeline Variables Reveal Clinically Relevant Subgroups. A) Principal component analysis of the 15 hemodynamic single beat parameters obtained from the computational pipeline. Data points are plotted based on their scores on the first two principal scores (PC1 and PC2). K-Means clustering identified two groups. For each group, an ellipse representing the covariance matrix is superimposed, where the tilt represents covariance and widths along the principal axes represent the standard deviations of PC1 and PC2. B) Heatmap of the 15 parameters in two clusters after normalization using Z-Score. C) Kaplan-Meier survival analysis with event free survival from diagnosis to combined events. The shaded bands indicate 95% confidence intervals. The number at risk table is shown at the bottom. Group A showed reduced event free survival compared to group B.

**Figure 4.**
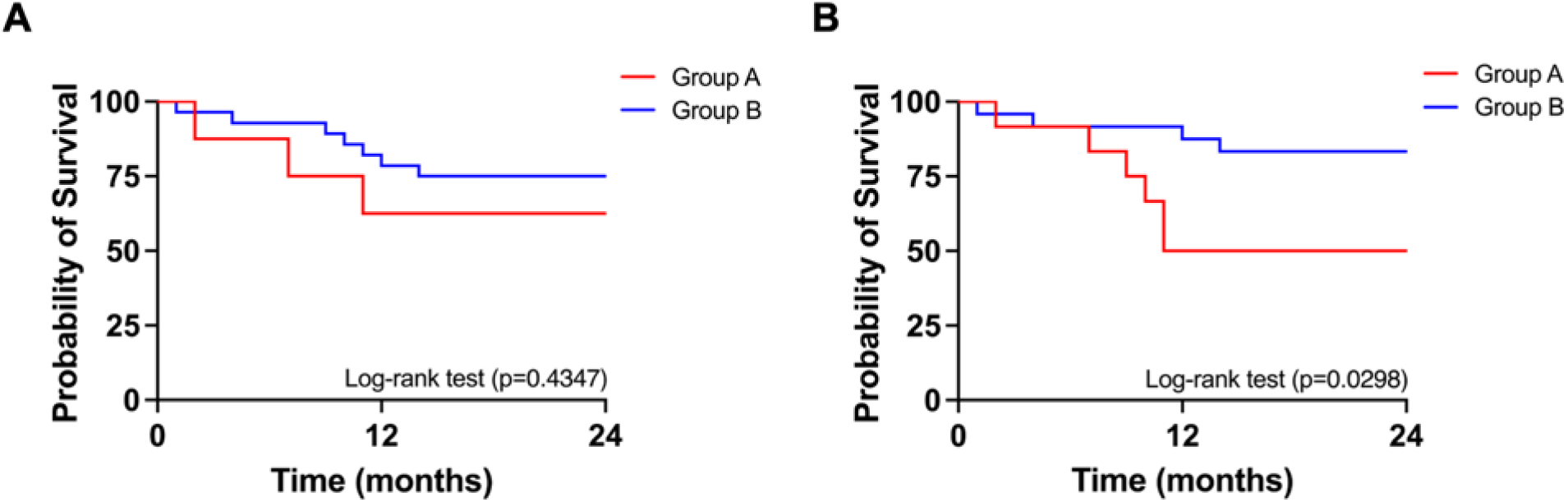
Inclusion of Single-Beat Variables Improves Phenotypic Clustering and Prognostic Separation. Kaplan–Meier curves for 24-month event-free survival in the two groups obtained when K-means clustering was applied to dataset consisting of routine clinical data *alone* (RHC and CMR imaging variables). The groups show no significant difference in survival (p=0.4347). B) Kaplan–Meier curves for groups generated after incorporating single_beat indices derived from computational pipeline with the same clinical variables. The new feature set yielded two groups with markedly different survival trajectories (p=0.0298).

**Table 3.**
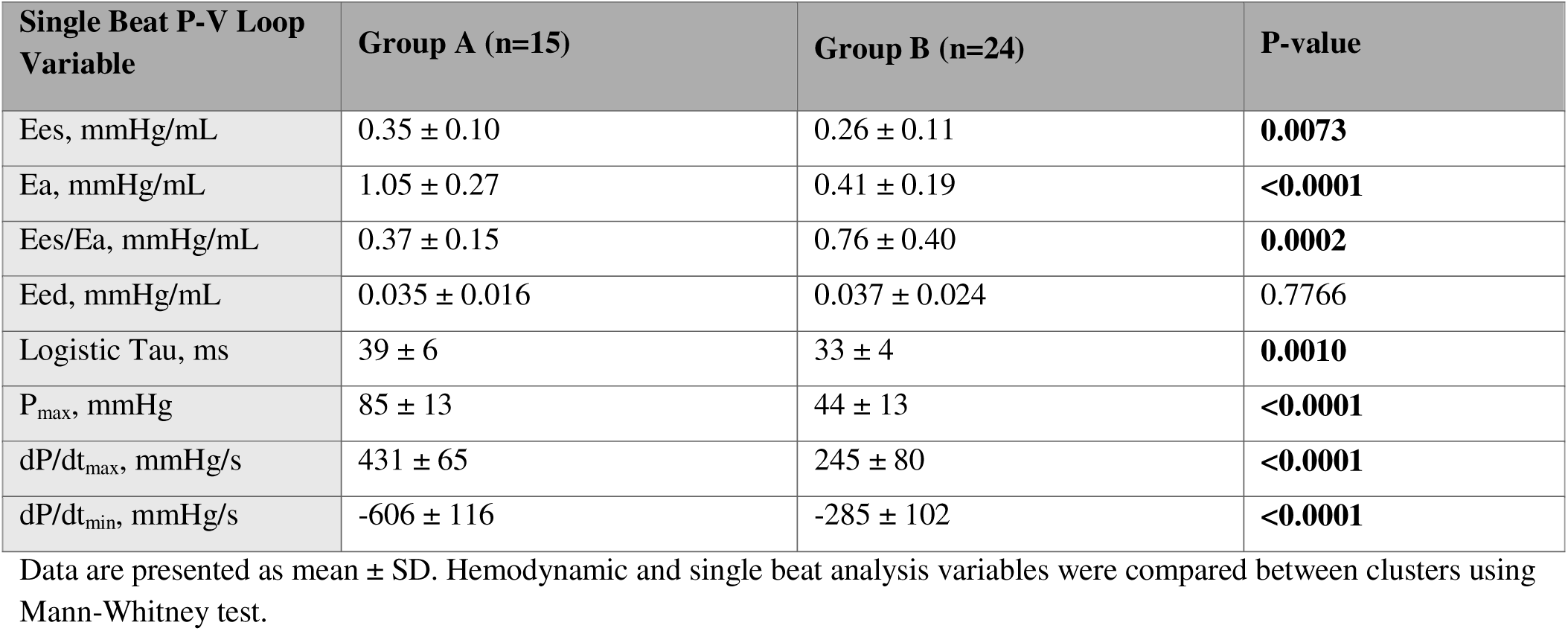
Comparisons for Single-Beat Analysis Derived Hemodynamic Characteristics between Subgroups.

**Table 4.**
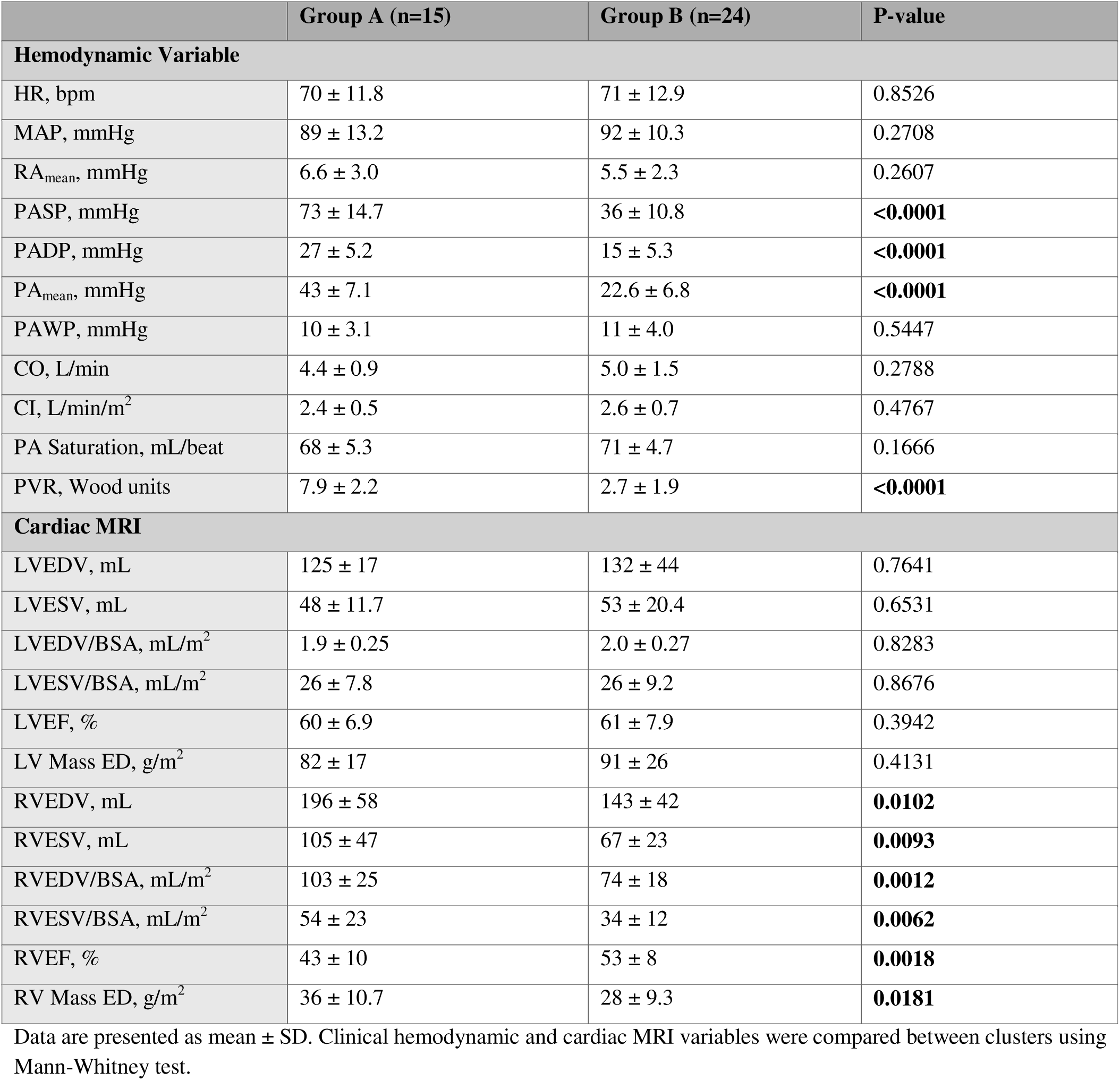
Comparisons for RHC-derived hemodynamic and Cardiac MRI variables between Subgroups.

|  | Group A (n=15) | Group B (n=24) | P-value |
| --- | --- | --- | --- |
| **Hemodynamic Variable** |  |  |  |
| HR, bpm | 70 ± 11.8 | 71 ± 12.9 | 0.8526 |
| MAP, mmHg | 89 ± 13.2 | 92 ± 10.3 | 0.2708 |
| RA<sub>mean</sub>, mmHg | 6.6 ± 3.0 | 5.5 ± 2.3 | 0.2607 |
| PASP, mmHg | 73 ± 14.7 | 36 ± 10.8 | **<0.0001** |
| PADP, mmHg | 27 ± 5.2 | 15 ± 5.3 | **<0.0001** |
| PA<sub>mean</sub>, mmHg | 43 ± 7.1 | 22.6 ± 6.8 | **<0.0001** |
| PAWP, mmHg | 10 ± 3.1 | 11 ± 4.0 | 0.5447 |
| CO, L/min | 4.4 ± 0.9 | 5.0 ± 1.5 | 0.2788 |
| CI, L/min/m<sup>2</sup> | 2.4 ± 0.5 | 2.6 ± 0.7 | 0.4767 |
| PA Saturation, mL/beat | 68 ± 5.3 | 71 ± 4.7 | 0.1666 |
| PVR, Wood units | 7.9 ± 2.2 | 2.7 ± 1.9 | **<0.0001** |
| **Cardiac MRI** |  |  |  |
| LVEDV, mL | 125 ± 17 | 132 ± 44 | 0.7641 |
| LVESV, mL | 48 ± 11.7 | 53 ± 20.4 | 0.6531 |
| LVEDV/BSA, mL/m<sup>2</sup> | 1.9 ± 0.25 | 2.0 ± 0.27 | 0.8283 |
| LVESV/BSA, mL/m<sup>2</sup> | 26 ± 7.8 | 26 ± 9.2 | 0.8676 |
| LVEF, % | 60 ± 6.9 | 61 ± 7.9 | 0.3942 |
| LV Mass ED, g/m<sup>2</sup> | 82 ± 17 | 91 ± 26 | 0.4131 |
| RVEDV, mL | 196 ± 58 | 143 ± 42 | **0.0102** |
| RVESV, mL | 105 ± 47 | 67 ± 23 | **0.0093** |
| RVEDV/BSA, mL/m<sup>2</sup> | 103 ± 25 | 74 ± 18 | **0.0012** |
| RVESV/BSA, mL/m<sup>2</sup> | 54 ± 23 | 34 ± 12 | **0.0062** |
| RVEF, % | 43 ± 10 | 53 ± 8 | **0.0018** |
| RV Mass ED, g/m<sup>2</sup> | 36 ± 10.7 | 28 ± 9.3 | **0.0181** |

Data are presented as mean ± SD. Clinical hemodynamic and cardiac MRI variables were compared between clusters using Mann-Whitney test.

## DISCUSSION

We present an accessible, easy-to-use computational pipeline program, available in the form of a downloadable application, for extraction of single-beat derived hemodynamics from a standard clinical RHC report. We demonstrate, in three independent cohorts with different PH etiologies, a strong agreement between our new methodology and the standard single beat estimate approach. Indices of RV function taken from our pipeline are prognostically significant, and subsequent cluster analysis performed using obtained indices identifies a PH patient subgroup with high afterload, uncoupled RVs, and worse outcomes.

To date, there are no publicly available tools, let alone clinically applicable tools, for single-beat RV analysis apart from custom scripts (e.g., MATLAB, R). Single-beat analysis is currently done by researchers using custom scripts. This poses significant barriers for clinicians or researchers without prior computational experience who may be interested in performing single-beat analysis to assess RV function in their patients. The pipeline we have developed here comes in the form of an easy-to-use graphical user interface (GUI) that does not require any prior programming knowledge. The input to our software is a screenshot of a standard clinical right heart catheterization and no additional hardware (e.g., analog pressure catheter, conductance catheter, etc.).

Our novel tool builds upon the methodology of the single beat approach. The fundamental principle behind single beat analysis, as developed by Sunagawa et al., involves fitting the isovolumic portions of the pressure-time tracing with a sinusoidal wave, which allows for estimation of the theoretical peak systolic pressure during isovolumic contraction, termed P_max_(27). End-systolic elastance is then estimated as the difference between P_max_ and end-systolic pressure divided by stroke volume. In the normal heart, this formulation appears to accurately reflect end-systolic elastance by multi-beat analysis(16,27). However, determinants of the accuracy of this method depend on the selection of the isovolumic ranges of the pressure waveform as well as the relative ratio of dP/dt_max_ to dP/dt_min_, as the use of a symmetric sinusoid constrains a ratio of 1, which may not be the case if marked diastolic dysfunction is present(28,29). Selection of the isovolumic portion involves calculation of either the first or second derivative of the RV pressure waveform, with the second derivative method more robust and widely used, also applied in our approach(17). The sinusoidal wave may not fit the pressure-time trace most commonly observed in waveforms from cardiomyopathy patients, resulting in an underestimation of P_max_. This is often the source of error for the calculation of Ees when compared to multi-beat analysis.(30) Our tool employs a novel yet simple piecewise sinusoid, which allows for differences in the relative magnitude of dP/dt_max_ to dP/dt_min_ while maintaining accurate fits with the isovolumic portions of the pressure waveform. In addition, our approach is completely analytic and does not rely upon nonlinear regression, yielding greater robustness. The only other alternative to this analysis avoids calculation of P_max_ entirely by leveraging the relatively preserved normalized time-varying elastance(23). While true for the LV, whether this principle applies to the RV is unknown.

Failure of the RV to increase contractility in the face of high afterload, as in decompensated PAH, is a primary driver of clinical outcomes, with several RV contractile measures derived from single- and multi-beat methods identified to be predictive of poor clinical outcomes(31,32). RV contractility is measured by end-systolic elastance, which in the normal ventricle increases as afterload (Ea) increases(33). RV-pulmonary arterial (RV-PA) coupling describes the concept that the RV will increase its contractility in response to afterload while minimizing energy loss (i.e., maximizing efficiency)(34). Indices of RV-PA coupling, be it as quantified by Ees/Ea or alternative methods (e.g., SV/ESV), provide superior prognostic information and better predict clinical outcomes compared to isolated measures of RV contractility or afterload alone(35–39). Cluster analysis of single-beat parameters obtained from our computational pipeline yielded two sub-phenotypes. Counterintuitively, the subgroup with worse outcomes had increased end-systolic elastance and P_max_. Their contractility was not itself predictive of poor outcomes but rather associated with an insufficient ability to counteract even higher afterload. As such, the Ees/Ea ratio was significantly worse in the poor outcome group. Thus, this points to this ratio as a key determinant of RV compensation and outcomes rather than focusing on RV contractility alone. This highlights again how important it would be to derive this ratio in the clinical setting rather than relying on isolated clinical indices of RV contractility.

The idea that RV-PA uncoupling is a significant prognostic indicator is not new. What is important to consider, however, are the current methods by which it is clinically assessed. The most easily obtained is the tricuspid annular systolic planar excursion (TAPSE) to PA systolic pressure ratio, measured by non-invasive methods from echocardiography(40,41). While demonstrating prognostic significance, the measurement of TAPSE is fraught with limitations. TAPSE is a linear measurement and depends on the angle of acquisition, is load-dependent and inaccurate when the RV is dilated, and is further limited by the fact that longitudinal contraction decreases early in the disease, while in later disease, radial contraction becomes more relevant.(40,42,43) Echo-derived PASP is also fraught with error, with up to 20 mmHg discordance from invasively measured PASP, even from the same day(44). These issues are particularly significant considering that the subgroup with the lowest Ees/Ea ratio in our study also presented with RV dilatation and hypertrophy on CMR. The pipeline presented here provides a robust alternative that removes many sources of measurement error. Its only downside is the requirement of an invasive measurement; with that said, serial RHC is already performed with regularity in patients with PH and HF, with minimal risk and complication. The procedure can be done not only in the catheterization laboratory but also is done routinely in the intensive care unit setting.

One notable observation is the presence of RV hypertrophy and diastolic dysfunction, as quantified by *τ*_logistic_, in the cluster subgroup with RV-PA uncoupling and poor outcomes. This highlights a correlation between reduced RV contractility and concomitant diastolic dysfunction. Recently, our group identified that, of all multi-beat PV loop indices, it was actually end-*diastolic* elastance that was most predictive of cardiac index reserve – superior even to the Ees/Ea ratio.(3) This leads to the intriguing hypothesis that RV diastolic dysfunction is an important driver of systolic reserve dysfunction in PH.(45) Given the importance of increased RV stiffness and RV function, measurement of Eed, or end-diastolic elastance, in PH patients may provide additional insight into RV hemodynamics.

### Limitations

The current study has several limitations. Digitization of waveforms from images is limited by pixel resolution and may result in loss of higher frequency signals and potential underestimation of dP/dt and Ees. In our study, up-sampling was performed to minimize the source of error with ∼2,000 pixels collected per second. Reports of the accuracy of single-beat analysis vis-à-vis multi-beat analysis are variable, and so the estimates we obtained may not reflect true contractility. We did attempt to rectify this by comparing our results to those from multi-beat analysis in our derivation cohort, identifying moderate-to-good concordance between the two. The small number of patients for outcomes analysis may have resulted in reduced power. Finally, our findings were limited to small cohorts, and although we sought to overcome this using two external validation cohorts, we would benefit from further optimization and validation in larger cohorts.

## CONCLUSION

We present a novel computational pipeline that only requires a screenshot from the electronic health record of an RV pressure-time tracing from a standard right heart catheterization to perform single-beat analysis for assessment of RV contractility. Our tool shows high accuracy in different sites, on different image formats, and in two groups of PH patients when compared to traditional single beat analysis. This is significant given that our agnostic cluster algorithm identifies the additive value of this analysis to other indices important for assessment of RV function in an RHC, showing that RV-PA uncoupling is a key discriminator of outcomes as is RV diastolic dysfunction. In summary, we present an easy-to-use, accessible tool for assessment of RV function, which may be able to detect RV failure before its end stages.

## Data Availability

All data produced in the present study are available upon reasonable request to the authors.

## Central Illustration

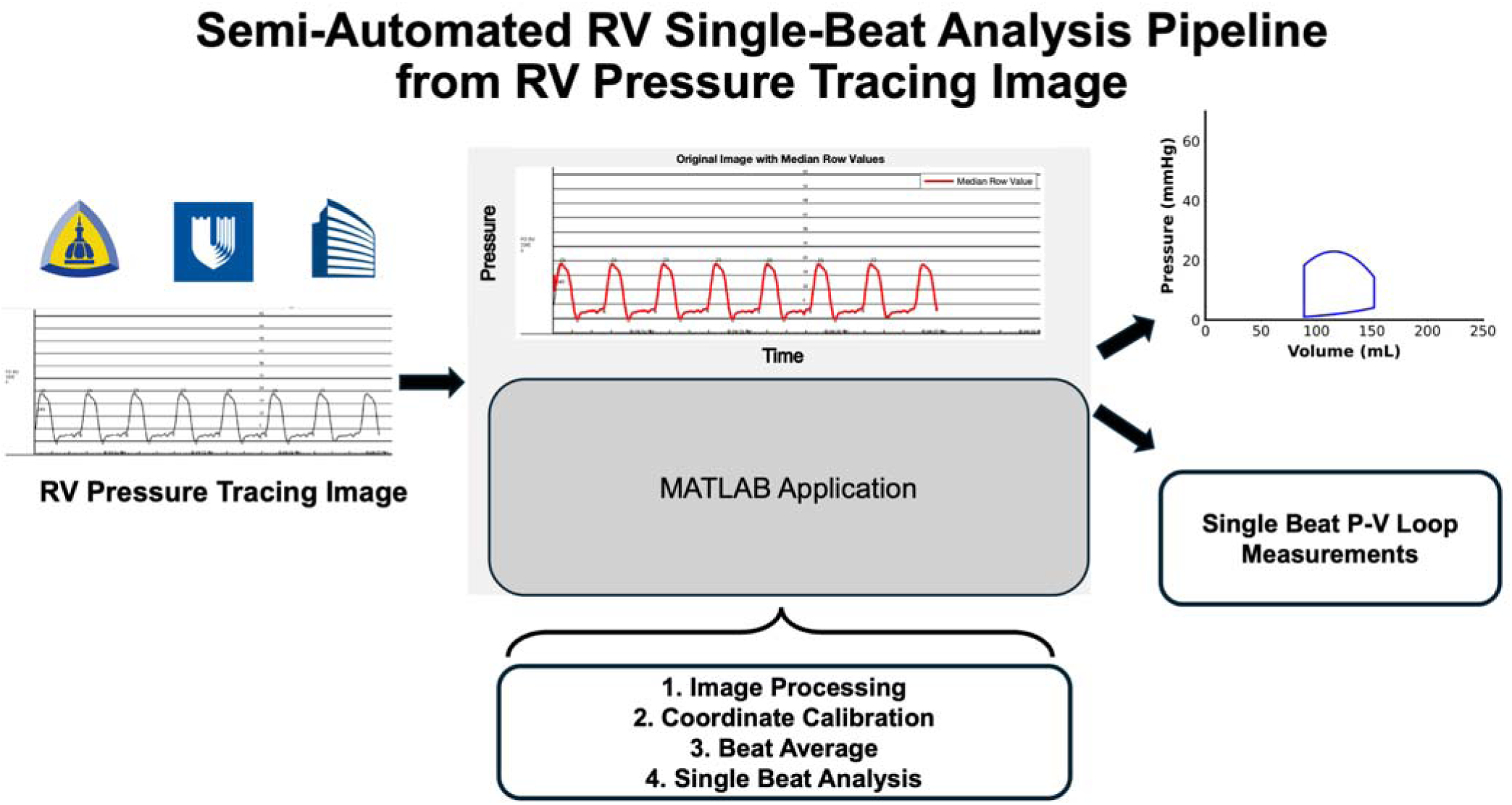
Central Illustration: A Semi-Automated RV Single-Beat Analysis Pipeline Can Successfully Generate Single Beat P-V Loop Parameters from an Image of RV Pressure Tracing Image. An application in MATLAB was developed to perform semi-automatic single-beat analysis and generation of right ventricular pressure-volume loop from an image of a right ventricular pressure tracing image and stroke volume from RHC.

## APPENDIX

Determination of peak isovolumic pressure (P_max_) is necessary for single beat analysis. Traditionally, P_max_ is obtained by fitting a sinusoid with dP/dt_max_ and dP/dt_min_ obtained from the numerical first derivative of the pressure waveform signal and fitting a sine wave from the pressure 20% of the maximum and minimum dP/dt. This approach however requires the ratio of dP/dt_max_ and dP/dt_min_ to be close to one. While reasonable for the normal heart, this presents effective application of single beat analysis in the save of severe diastolic dysfunction. To address this, we developed a piecewise sinusoidal approach for determination of P_,max._

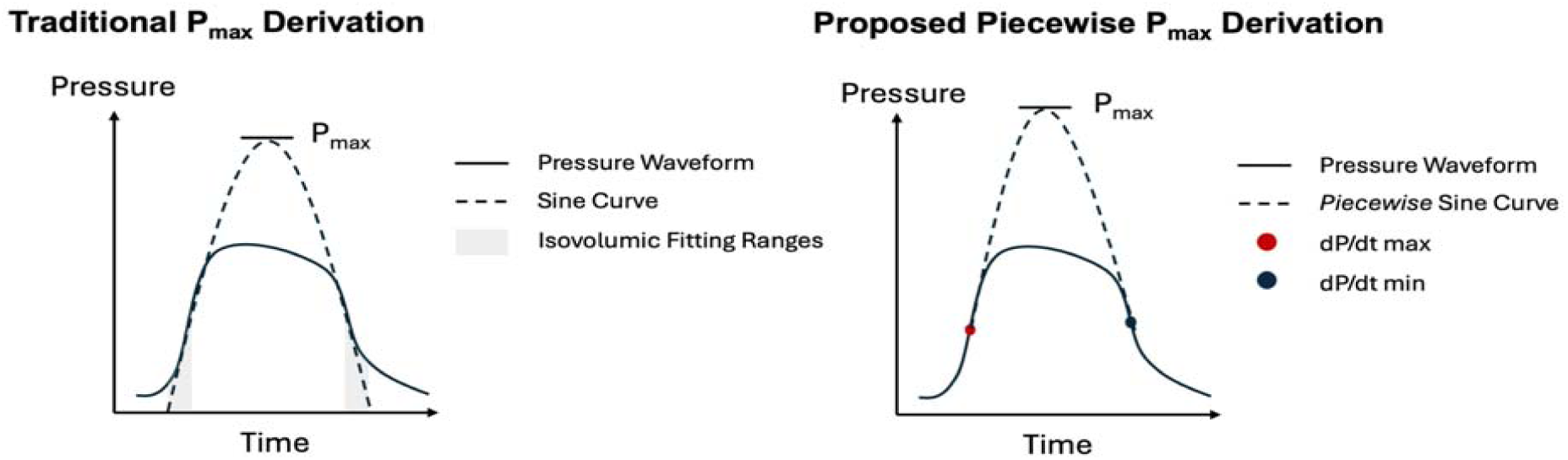

Let *T* be the absolute time scale. To obtain P_max_ we take the pressure waveform and fit it to the equation. We parameterize time such that *t* = *T* – *t_dP/dt max_*, where *t_dP/dt max_* is the time at which the first derivative is maximum. Similarly, we define *t*\* = (*t_dP/dt min_* – *t_dP/dt max_*) – *t* = *T_p_* – *t*, where *t_dP/dt min_* is the time at which the first derivative of pressure is minimum. We define a piecewise sinusoid P(t) as

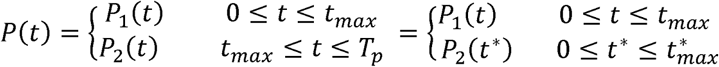

Where 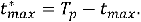 From the pressure waveform, we know the pressure at t = 0 and t*= 0 is the amplitude of the pressure waveform at *t_dP/dt max_* and *t_dP/dt min_*, respectively, which we denote *P_dP/dt max_* and *P_dP/dt min_*. From this condition, we write the piecewise sine function as

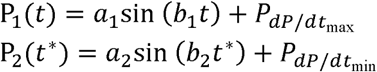

We also know that at t = 0 and t*= 0, the derivative of our piecewise sinusoid must be either dP/dt_max_ or dP/dt_min_, or

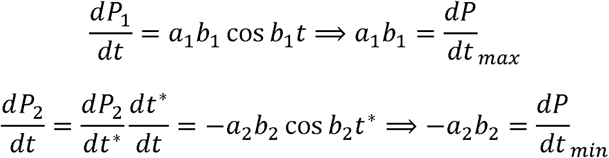

Finally, we enforce that when P = P_max_, the piecewise sinusoid is continuous and smooth. We enforce the smoothness constraint by forcing each sinusoid to be at its maximum at P_max_, or dP_1_/dt = dP_2_/dt = 0. With these conditions, we can write the final three equations. For continuity, we have

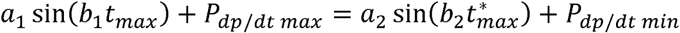

For smoothness we have:

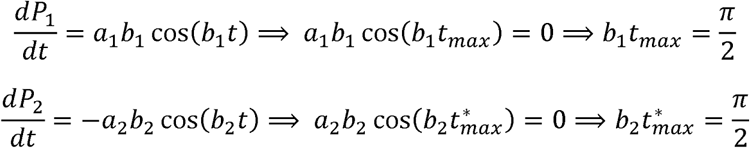

We thus have a series of 7 equations for which we can obtain the piecewise sinusoidal to obtain P_max_.

## ABBREVIATIONS

6MWD: 6-Minute Walk Distance
BMI: Body Mass Index
CI: Cardiac Index
CO: Cardiac Output
ESC/ERS: European Society of Cardiology/European Respiratory Society
HFpEF: Heart Failure with Preserved Ejection Fraction
mPAP: mean Pulmonary Arterial Pressure
NT-proBNP: NT-pro-Brain Natriuretic Peptide
PAH: Pulmonary Arterial Hypertension
PAWP: Pulmonary Artery Wedge Pressure
PDE-5i: Phosphodiesterase Type 5 inhibitor
PH: Pulmonary Hypertension
PAH: Pulmonary Arterial Hypertension
PVR: Pulmonary Vascular Resistance
RA: Right Atrium
RV: Right Ventricle
RHC: Right Heart Catheterization
SD: Standard Deviation
WHO: World Health Organization
WU: Wood Units

## REFERENCES

1. Naeije R, Manes A. The right ventricle in pulmonary arterial hypertension. European respiratory review 2014;23:476–487.

2. Sanz J, Sánchez-Quintana D, Bossone E, Bogaard HJ, Naeije R. Anatomy, function, and dysfunction of the right ventricle: JACC state-of-the-art review. Journal of the American College of Cardiology 2019;73:1463–1482.

3. Salazar IMC, Lancaster AC, Jani VP et al. Poor Cardiac Output Reserve in Pulmonary Arterial Hypertension is Associated With Right Ventricular Stiffness and Impaired Interventricular Dependence. European Respiratory Journal 2024.

4. Handoko M, De Man F, Allaart C, Paulus W, Westerhof N, Vonk-Noordegraaf A. Perspectives on novel therapeutic strategies for right heart failure in pulmonary arterial hypertension: lessons from the left heart. European Respiratory Review 2010;19:72–82.

5. Houston BA, Brittain EL, Tedford RJ. Right ventricular failure. New England Journal of Medicine 2023;388:1111–1125.

6. Takeda K, Takayama H, Colombo PC et al. Incidence and clinical significance of late right heart failure during continuous-flow left ventricular assist device support. The Journal of Heart and Lung Transplantation 2015;34:1024–1032.

7. Rame JE, Pagani FD, Kiernan MS et al. Evolution of late right heart failure with left ventricular assist devices and association with outcomes. Journal of the American College of Cardiology 2021;78:2294–2308.

8. Lampert BC, Teuteberg JJ. Right ventricular failure after left ventricular assist devices. The Journal of Heart and Lung Transplantation 2015;34:1123–1130.

9. Mehra MR, Goldstein DJ, Cleveland JC et al. Five-year outcomes in patients with fully magnetically levitated vs axial-flow left ventricular assist devices in the MOMENTUM 3 randomized trial. Jama 2022;328:1233–1242.

10. Houston BA, Shah KB, Mehra MR, Tedford RJ. A new “twist” on right heart failure with left ventricular assist systems. Elsevier, 2017:701–707.

11. Tedford RJ, Mudd JO, Girgis RE et al. Right ventricular dysfunction in systemic sclerosis–associated pulmonary arterial hypertension. Circulation: Heart Failure 2013;6:953–963.

12. Aslam MI, Jani V, Lin B et al. Pulmonary artery pulsatility index predicts right ventricular myofilament dysfunction in advanced human heart failure. European journal of heart failure 2021;23:339.

13. Frankfurter C, Molinero M, Vishram-Nielsen JK et al. Predicting the risk of right ventricular failure in patients undergoing left ventricular assist device implantation: a systematic review. Circulation: Heart Failure 2020;13:e006994.

14. Konstam MA, Kiernan MS, Bernstein D et al. Evaluation and management of right-sided heart failure: a scientific statement from the American Heart Association. Circulation 2018;137:e578–e622.

15. He Q, Lin Y, Zhu Y et al. Clinical usefulness of right ventricle–pulmonary artery coupling in cardiovascular disease. Journal of Clinical Medicine 2023;12:2526.

16. Brimioulle S, Wauthy P, Ewalenko P et al. Single-beat estimation of right ventricular end-systolic pressure-volume relationship. American Journal of Physiology-Heart and Circulatory Physiology 2003;284:H1625–H1630.

17. Heerdt PM, Kheyfets V, Charania S, Elassal A, Singh I. A pressure-based single beat method for estimation of right ventricular ejection fraction: proof of concept. European Respiratory Journal 2020;55.

18. Hassan HJ, Naranjo M, Ayoub N et al. Improved survival for patients with systemic sclerosis–associated pulmonary arterial hypertension: the Johns Hopkins Registry. American journal of respiratory and critical care medicine 2023;207:312–322.

19. Simonneau G, Montani D, Celermajer DS et al. Haemodynamic definitions and updated clinical classification of pulmonary hypertension. European respiratory journal 2019;53.

20. Van Den Hoogen F, Khanna D, Fransen J et al. 2013 classification criteria for systemic sclerosis: an American College of Rheumatology/European League against Rheumatism collaborative initiative. Arthritis & Rheumatism 2013;65:2737–2747.

21. Masi AT, Diagnostic SFSCotARA, Committee TC. Preliminary criteria for the classification of systemic sclerosis (scleroderma). Arthritis & Rheumatism 1980;23:581–590.

22. Hsu S, Houston BA, Tampakakis E et al. Right ventricular functional reserve in pulmonary arterial hypertension. Circulation 2016;133:2413–2422.

23. Senzaki H, Chen C-H, Kass DA. Single-beat estimation of end-systolic pressure-volume relation in humans: a new method with the potential for noninvasive application. Circulation 1996;94:2497–2506.

24. Ogilvie LM, Edgett BA, Huber JS et al. Hemodynamic assessment of diastolic function for experimental models. American Journal of Physiology-Heart and Circulatory Physiology 2020;318:H1139–H1158.

25. Klotz S, Dickstein ML, Burkhoff D. A computational method of prediction of the end-diastolic pressure– volume relationship by single beat. Nature protocols 2007;2:2152–2158.

26. Chen C-H, Fetics B, Nevo E et al. Noninvasive single-beat determination of left ventricular end-systolic elastance in humans. Journal of the American College of Cardiology 2001;38:2028–2034.

27. Sunagawa K, Yamada A, Senda Y et al. Estimation of the hydromotive source pressure from ejecting beats of the left ventricle. IEEE Transactions on Biomedical Engineering 1980:299–305.

28. Misbach GA, Glantz SA. Changes in the diastolic pressure-diameter relation after ventricular function curves. American Journal of Physiology-Heart and Circulatory Physiology 1979;237:H644–H648.

29. Kass DA. Assessment of diastolic dysfunction: invasive modalities. Cardiology clinics 2000;18:571–586.

30. Pak PH, Maughan WL, Baughman KL, Kass DA. Marked discordance between dynamic and passive diastolic pressure-volume relations in idiopathic hypertrophic cardiomyopathy. Circulation 1996;94:52–60.

31. Vonk-Noordegraaf A, Haddad F, Chin KM et al. Right heart adaptation to pulmonary arterial hypertension: physiology and pathobiology. Journal of the American College of Cardiology 2013;62:D22–D33.

32. Fine NM, Chen L, Bastiansen PM et al. Outcome prediction by quantitative right ventricular function assessment in 575 subjects evaluated for pulmonary hypertension. Circulation: Cardiovascular Imaging 2013;6:711–721.

33. Hsu S, Fang JC, Borlaug BA. Hemodynamics for the heart failure clinician: a state-of-the-art review. Journal of cardiac failure 2022;28:133–148.

34. Kass DA, Beyar R. Evaluation of contractile state by maximal ventricular power divided by the square of end-diastolic volume. Circulation 1991;84:1698–1708.

35. Wu Y, Tian P, Liang L et al. Combined use of right ventricular coupling and pulmonary arterial elastance as a comprehensive stratification approach for right ventricular function. Clinical and Translational Science 2023;16:1582–1593.

36. Jone P-N, Schäfer M, Pan Z, Ivy DD. Right ventricular-arterial coupling ratio derived from 3-dimensional echocardiography predicts outcomes in pediatric pulmonary hypertension. Circulation: Cardiovascular Imaging 2019;12:e008176.

37. Jung Y-H, Ren X, Suffredini G, Dodd-o JM, Gao WD. Right ventricular diastolic dysfunction and failure: a review. Heart Failure Reviews 2022;27:1077–1090.

38. Vonk Noordegraaf A, Westerhof BE, Westerhof N. The relationship between the right ventricle and its load in pulmonary hypertension. Journal of the American College of Cardiology 2017;69:236–243.

39. Vanderpool RR, Pinsky MR, Naeije R et al. RV-pulmonary arterial coupling predicts outcome in patients referred for pulmonary hypertension. Heart 2015;101:37–43.

40. Tello K, Axmann J, Ghofrani HA et al. Relevance of the TAPSE/PASP ratio in pulmonary arterial hypertension. International journal of cardiology 2018;266:229–235.

41. Schmeisser A, Rauwolf T, Groscheck T et al. Pressure–volume loop validation of TAPSE/PASP for right ventricular arterial coupling in heart failure with pulmonary hypertension. European Heart Journal-Cardiovascular Imaging 2021;22:168–176.

42. Aloia E, Cameli M, D’Ascenzi F, Sciaccaluga C, Mondillo S. TAPSE: an old but useful tool in different diseases. International journal of cardiology 2016;225:177–183.

43. Guazzi M. Use of TAPSE/PASP ratio in pulmonary arterial hypertension: an easy shortcut in a congested road. International Journal of Cardiology 2018;266:242–244.

44. Fisher MR, Forfia PR, Chamera E et al. Accuracy of Doppler echocardiography in the hemodynamic assessment of pulmonary hypertension. American journal of respiratory and critical care medicine 2009;179:615–621.

45. Trip P, Rain S, Handoko ML et al. Clinical relevance of right ventricular diastolic stiffness in pulmonary hypertension. European Respiratory Journal 2015;45:1603–1612.

