## Supplemental Files for "Development of Computational Pipeline for Right Ventricular Hemodynamic Single-Beat Analysis"

**SUPPLEMENTAL TABLES**

**Supplemental Table 1. Baseline Demographics, Clinical Characteristics, and Haemodynamic Variables.** Data are presented as n (%), mean (SD), and median (IQR). PH = Pulmonary Hypertension, Ssc-PAH = systemic sclerosis-associated PAH, SSc-ILD-PH = SSc-PH related to interstitial lung disease, SSc-HFpEF = systemic sclerosis-associated heart failure with preserved ejection fraction, ERA = endothelin receptor agonist, PDE5i = phosphodiesterase 5 inhibitor, HR = heart rate, MAP = mean arterial pressure, RAP = right atrial pressure, PASP = pulmonary arterial systolic pressure, PADP = pulmonary arterial diastolic pressure, PA_mean_ = mean pulmonary artery pressure, PAWP = pulmonary artery wedge pressure, CO = cardiac output, CI = cardiac index, PVR = pulmonary vascular resistance.

| **Johns Hopkins Cohort**  **Baseline demographics/characteristics** | **Overall (n=39)** |
| --- | --- |
| **Age, y** | 57 (12) |
| **Female Sex, %** | 36 (92.3) |
| **Race, %** |  |
| White | 27 (69.2) |
| Black | 9 (23.1) |
| Asian | 2 (5.1) |
| Hispanic | 1 (2.6) |
| **Height, m** | 1.65 (0.08) |
| **Weight, kg** | 82 (19) |
| **BSA, m^2^** | 1.9 (0.3) |
| **BMI, kg/m^2^** | 30 (6) |
| **NYHA Class** |  |
| 1 | 5 (12.8) |
| 2 | 26 (66.7) |
| 3 | 8 (20.5) |
| **Subgroup, %** |  |
| No PH | 10 (25.6) |
| iPAH | 12 (30.8) |
| SSc-PAH | 12 (30.8) |
| SSc-ILD-PH | 1 (2.6) |
| SSc-HFpEF | 3 (7.7) |
| SSc-Other | 1 (2.6) |
| **Diagnosis at time of study, count, %** |  |
| Incident | 8 (20.5) |
| Prevalent | 22 (56.4) |
| N/A | 9 (23.1) |
| **W.H.O. Functional Class, count, %** |  |
| 0 | 11 (28.2) |
| 1 | 24 (61.5) |
| 2 | 3 (7.7) |
| 3 | 1 (2.6) |
| **SSc Subtype, %** |  |
| None | 13 (33.3) |
| Limited | 11 (28.2) |
| Diffuse | 5 (12.8) |
| Other | 10 (25.6) |
| **Medication, %** |  |
| ERA | 17 (43.6) |
| PDE5i | 19 (48.7) |
| Prostacyclin | 2 (5.1) |
| Monotherapy | 5 (12.8) |
| Dual therapy | 15 (38.5) |
| Triple therapy | 1 (2.6) |
| No therapy | 18 (46.2) |
| Calcium Channel Blockers (CCB) | 14 (35.9) |
| Aldosterone Receptor Antagonists (MRAs) | 4 (10.3) |
| Loop Diuretic | 18 (46.2) |
| **6 Minute Walk Test, m** | 414 (110) |
| **Creatinine, mg/dL** | 0.90 (0.24) |
| **Hemoglobin, g/dL** | 12.6 (1.9) |
| **NT-proBNP, pg/mL** | 222 (136-397) |
| **Haemodynamic Variable** | |
| HR, bpm | 69 (62-78) |
| MAP, mmHg | 91 (12) |
| RAP, mmHg | 6 (2.6) |
| PASP, mmHg | 51 (22) |
| PADP, mmHg | 20 (8) |
| PA_mean_, mmHg | 31 (12) |
| PAWP, mmHg | 10 (4) |
| CO, L/min | 4.8 (1.4) |
| CI, L/min/m^2^ | 2.5 (0.6) |
| PA Saturation, mL/beat | 70 (5) |
| PVR, Wood units | 5 (3) |
| **Medical University of South Carolina**  **Baseline demographics/characteristics** | **Overall (n=17)** |
| **Age, y** | 62 (12) |
| **Female Sex, %** | 11 (64.7) |
| **Race, %** |  |
| White | 15 (88.2) |
| Black | 2 (11.8) |
| Asian | 0 (0.0) |
| Hispanic | 0 (0.0) |
| **BMI, kg/m^2^** | 32 (6) |
| **Echo EF, %** | 68 (6) |
| **CO, L/min** | 5.4 (1.1) |
| **Duke University**  **Baseline demographics/characteristics** | **Overall (n=20)** |
| **Age, y** | 65 (12) |
| **Female Sex, %** | 6 (30.0) |
| **Race, %** |  |
| White | 10 (50.0) |
| Black | 9 (45.0) |
| Asian | 1 (5.0) |
| Hispanic | 0 (0.0) |
| **BMI, kg/m^2^** | 30 (8) |
| **Echo EF, %** | 29 (12) |
| **CO, L/min** | 4.9 (1.4) |

**Supplemental Table 2. Goodness of Fit Metrics between Gold Standard Single Beat P-V Loop Variables and Pipeline Results in 3 Centers.** Data are presented as mean ± SD. GS = Gold standard, CCC = Lin's concordance correlation coefficient, B = Beta coefficient, R = Pearson correlation coefficient, Ees = end systolic elastance, Ea = arterial elastance, Ees/Ea = RV-PA coupling, Eed = end diastolic elastance, RVEF = right ventricular ejection fraction, RVEDV = right ventricular end diastolic volume, RVESV = right ventricular end systolic volume, RVEDP = right ventricular end diastolic pressure, RVESP = right ventricular end systolic pressure

| **Single Beat P-V Loop Variable** | **Mean (GS)** | **Mean (Pipeline)** | **CCC** | **B** | **R** | **Mean Difference** |
| --- | --- | --- | --- | --- | --- | --- |
| Johns Hopkins | | | | | | |
| Ees, mmHg/mL | 0.35 ± 0.12 | 0.3 ± 0.1 | 0.58 | 0.82 | 0.96 | -0.056 ± 0.097 |
| Ea, mmHg/mL | 0.70 ± 0.46 | 0.65 ± 0.38 | 0.88 | 0.88 | 0.97 | -0.04 ± 0.20 |
| Ees/Ea, mmHg/mL | 0.74 ± 0.47 | 0.61 ± 0.38 | 0.71 | 0.76 | 0.93 | -0.13 ± 0.31 |
| Eed, mmHg/mL | 0.04 ± 0.02 | 0.036 ± 0.021 | 0.47 | 0.74 | 0.87 | -0.007 ± 0.023 |
| Logistic Tau, ms | 30 ± 5 | 35 ± 6 | 0.29 | 1.16 | 0.98 | 5.3 ± 5.9 |
| RVEF % | 38.9 ± 14.2 | 35 ± 12 | 0.73 | 0.85 | 0.97 | -3.8 ± 9.2 |
| RVEDV, mL | 208 ± 136 | 213 ± 73 | 0.33 | 0.79 | 0.86 | 4.6 ± 126.3 |
| RVESV, mL | 139 ± 134 | 144 ± 69 | 0.30 | 0.63 | 0.76 | 4.6 ± 126.3 |
| RVEDP, mmHg | 9 ± 10 | 6.9 ± 3.5 | 0.16 | 0.41 | 0.70 | -2.2 ± 9.3 |
| RVESP, mmHg | 44 ± 27 | 41 ± 21 | 0.85 | 0.86 | 0.97 | -3.1 ± 13.2 |
| P_max_, mmHg | 67 ± 28 | 60 ± 24 | 0.88 | 0.88 | 0.99 | -6.7 ± 11.3 |
| dP/dt_max_, mmHg/s | 430 ± 183 | 317 ± 118 | 0.37 | 0.68 | 0.93 | -113.4 ± 158.5 |
| dP/dt_min_, mmHg/s | -459 ± 321 | -408 ± 191 | 0.60 | 0.74 | 0.91 | 51.1 ± 231.4 |
| β | 0.027 ± 0.015 | 0.026 ± 0.013 | 0.51 | 0.85 | 0.90 | -0.001 ± 0.014 |
| MUSC | | | | | | |
| Ees, mmHg/mL | 0.26 ± 0.09 | 0.23 ± 0.09 | 0.78 | 0.88 | 0.98 | -0.03 ± 0.05 |
| Ea, mmHg/mL | 0.32 ± 0.14 | 0.36 ± 0.17 | 0.87 | 1.12 | 0.99 | 0.04 ± 0.07 |
| Ees/Ea, mmHg/mL | 0.88 ± 0.30 | 0.73 ± 0.30 | 0.72 | 0.83 | 0.98 | -0.15 ± 0.18 |
| Eed, mmHg/mL | 0.07 ± 0.02 | 0.032 ± 0.017 | 0.14 | 0.45 | 0.90 | -0.04 ± 0.02 |
| Logistic Tau, ms | 27 ± 4.1 | 24 ± 13 | 0.23 | 0.91 | 0.91 | -2.7 ± 11.6 |
| RVEF % | 45 ± 9 | 40 ± 11 | 0.73 | 0.89 | 0.99 | -5 ± 6 |
| RVEDV, mL | 179 ± 67 | 204 ± 71 | 0.83 | 1.12 | 0.99 | 25 ± 33 |
| RVESV, mL | 102 ± 53 | 127 ± 62 | 0.76 | 1.20 | 0.97 | 25 ± 33 |
| RVEDP, mmHg | 11 ± 3 | 6.3 ± 3.0 | 0.24 | 0.55 | 0.93 | -5.1 ± 3.1 |
| RVESP, mmHg | 23 ± 8 | 26 ± 11 | 0.80 | 1.13 | 0.98 | 2.9 ± 5.4 |
| P_max_, mmHg | 42 ± 10 | 43 ± 12 | 0.81 | 1.02 | 0.99 | 0.9 ± 6.8 |
| dP/dt_max_, mmHg/s | 262 ± 75 | 259 ± 73 | 0.30 | 0.94 | 0.95 | -3 ± 87 |
| dP/dt_min_, mmHg/s | -229 ± 73 | -262 ± 115 | 0.66 | 1.15 | 0.97 | -33 ± 74 |
| β | 0.031 ± 0.008 | 0.02 ± 0.01 | 0.10 | 0.61 | 0.86 | -0.01 ± 0.01 |
| Duke | | | | | | |
| Ees, mmHg/mL | 0.26 ± 0.16 | 0.17 ± 0.10 | 0.66 | 0.63 | 0.98 | -0.088 ± 0.083 |
| Ea, mmHg/mL | 0.73 ± 0.39 | 0.75 ± 0.40 | 0.99 | 1.02 | 1.00 | 0.015 ± 0.066 |
| Ees/Ea, mmHg/mL | 0.41 ± 0.20 | 0.30 ± 0.25 | 0.73 | 0.79 | 0.93 | -0.11 ± 0.14 |
| Eed, mmHg/mL | 0.04 ± 0.03 | 0.029 ± 0.013 | 0.29 | 0.53 | 0.87 | -0.014 ± 0.026 |
| Logistic Tau, ms | 30 ± 5.5 | 35 ± 8 | 0.39 | 1.14 | 0.98 | 4.6 ± 7.3 |
| RVEF % | 28 ± 9 | 21 ± 11 | 0.70 | 0.79 | 0.97 | -6.7 ± 5.0 |
| RVEDV, mL | 230 ± 81 | 339 ± 174 | 0.56 | 1.52 | 0.98 | 109 ± 105 |
| RVESV, mL | 169 ± 69 | 278 ± 164 | 0.47 | 1.71 | 0.97 | 109 ± 105 |
| RVEDP, mmHg | 9 ± 4 | 9.1 ± 4.5 | 0.85 | 1.00 | 0.97 | 0.1 ± 2.3 |
| RVESP, mmHg | 39 ± 16 | 40 ± 15 | 0.97 | 1.01 | 1.00 | 0.7 ± 3.5 |
| P_max_, mmHg | 53 ± 19 | 49 ± 16 | 0.94 | 0.92 | 1.00 | -3.9 ± 4.5 |
| dP/dt_max_, mmHg/s | 347 ± 174 | 277 ± 125 | 0.69 | 0.76 | 0.97 | -70 ± 104 |
| dP/dt_min_, mmHg/s | -331 ± 121 | -294 ± 116 | 0.89 | 0.89 | 0.99 | 37 ± 41 |
| β | 0.037 ± 0.020 | 0.036 ± 0.016 | 0.88 | 0.93 | 0.98 | -0.001 ± 0.009 |

##### Supplemental Table 3. Relationship between Single Beat Parameters from Pipeline and Outcome Variables as Standardized Hazard Ratio (95% CI). Data are presented as standardized hazard ratio (95% CI). Ees = end systolic elastance, Ea = arterial elastance, Ees/Ea = RV-PA coupling, Eed = end diastolic elastance, RVEF = right ventricular ejection fraction, RVEDV = right ventricular end diastolic volume, RVESV = right ventricular end systolic volume, RVEDP = right ventricular end diastolic pressure, RVESP = right ventricular end systolic pressure

| **Single Beat P-V Loop Variable** | **Death** | **P-val** | **HF Hosp** | **P-val** | **PAH Esc** | **P-val** | **Combined** | **P-val** |
| --- | --- | --- | --- | --- | --- | --- | --- | --- |
| Gold Standard | | | | | | | | |
| Ees, mmHg/mL | 0.93 (0.34-2.56) | 0.894 | 1.19 (0.47-2.97) | 0.714 | 1.24 (0.47-3.22) | 0.665 | 0.78 (0.46-1.33) | 0.360 |
| Ea, mmHg/mL | **8.71 (1.04-73.09)** | **0.046** | 3.74 (1.00-14.04) | 0.051 | 3.94 (0.99-15.75) | 0.052 | **2.50 (1.20-5.17)** | **0.014** |
| Ees/Ea, mmHg/mL | **0.22 (0.07-0.75)** | **0.016** | **0.32 (0.11-0.91)** | **0.033** | **0.26 (0.07-0.88)** | **0.031** | **0.33 (0.18-0.61)** | **0.000** |
| Eed, mmHg/mL | 4.06 (0.86-19.16) | 0.076 | 0.70 (0.29-1.66) | 0.415 | 1.22 (0.46-3.19) | 0.692 | 1.18 (0.64-2.19) | 0.599 |
| Logistic Tau, ms | 0.84 (0.34-2.08) | 0.708 | 0.57 (0.21-1.60) | 0.288 | 1.34 (0.53-3.43) | 0.537 | 0.94 (0.50-1.76) | 0.836 |
| RVEF % | **0.20 (0.04-0.93)** | **0.041** | **0.29 (0.08-0.99)** | **0.049** | **0.26 (0.07-0.94)** | **0.039** | **0.31 (0.15-0.63)** | **0.001** |
| RVEDV, mL | 4.45 (0.74-26.73) | 0.103 | 1.39 (0.54-3.59) | 0.495 | **8.73 (1.39-54.88)** | **0.021** | **1.96 (1.30-2.96)** | **0.001** |
| RVESV, mL | 4.94 (0.71-34.35) | 0.107 | 1.57 (0.71-3.48) | 0.269 | **9.68 (1.67-56.13)** | **0.011** | **1.99 (1.33-2.99)** | **0.001** |
| RVEDP, mmHg | **9.00 (1.74-46.48)** | **0.009** | 0.47 (0.04-5.79) | 0.558 | **5.74 (1.27-26.03)** | **0.024** | **1.88 (1.24-2.86)** | **0.003** |
| RVESP, mmHg | **5.34 (1.68-16.93)** | **0.004** | 1.97 (0.76-5.08) | 0.160 | **3.75 (1.29-10.93)** | **0.016** | **2.34 (1.33-4.09)** | **0.003** |
| P_max_, mmHg | **5.00 (1.63-15.30)** | **0.005** | 1.67 (0.63-4.46) | 0.305 | **3.47 (1.20-10.01)** | **0.021** | **2.03 (1.14-3.62)** | **0.016** |
| dP/dt_max_, mmHg/s | **3.93 (1.16-13.33)** | **0.028** | 1.12 (0.35-3.57) | 0.854 | 3.03 (0.73-12.62) | 0.127 | 1.54 (0.91-2.63) | 0.109 |
| dP/dt_min_, mmHg/s | **0.26 (0.09-0.74)** | **0.012** | **0.40 (0.18-0.93)** | **0.034** | **0.07 (0.01-0.75)** | **0.027** | **0.42 (0.26-0.68)** | **0.000** |
| β | 0.88 (0.45-1.69) | 0.696 | 1.10 (0.62-1.94) | 0.742 | 1.02 (0.57-1.80) | 0.953 | 1.04 (0.73-1.47) | 0.842 |
| Pipeline | | | | | | | | |
| Ees, mmHg/mL | 1.97 (0.62-6.22) | 0.250 | 1.42 (0.57-3.54) | 0.453 | 1.83 (0.71-4.76) | 0.214 | 1.36 (0.78-2.39) | 0.280 |
| Ea, mmHg/mL | 2.54 (0.66-9.76) | 0.175 | **5.86 (1.27-27.00)** | **0.023** | **4.49 (1.06-19.00)** | **0.041** | **2.09 (1.04-4.20)** | **0.038** |
| Ees/Ea, mmHg/mL | 0.62 (0.21-1.78) | 0.371 | **0.27 (0.08-0.87)** | **0.028** | 0.44 (0.16-1.18) | 0.102 | 0.59 (0.33-1.07) | 0.080 |
| Eed, mmHg/mL | 0.93 (0.29-2.97) | 0.902 | 1.48 (0.51-4.28) | 0.466 | 1.63 (0.66-4.04) | 0.293 | 1.31 (0.74-2.33) | 0.352 |
| Logistic Tau, ms | 1.18 (0.52-2.69) | 0.692 | 1.43 (0.77-2.64) | 0.260 | 1.31 (0.66-2.63) | 0.441 | 1.05 (0.65-1.70) | 0.827 |
| RVEF % | 0.54 (0.16-1.78) | 0.311 | **0.23 (0.06-0.92)** | **0.038** | 0.39 (0.13-1.22) | 0.105 | 0.57 (0.30-1.08) | 0.085 |
| RVEDV, mL | 1.27 (0.46-3.48) | 0.647 | 1.63 (0.67-4.00) | 0.282 | 1.92 (0.84-4.39) | 0.120 | 1.40 (0.83-2.39) | 0.209 |
| RVESV, mL | 1.33 (0.51-3.48) | 0.558 | 1.99 (0.87-4.57) | 0.105 | 1.96 (0.91-4.23) | 0.085 | 1.50 (0.91-2.47) | 0.114 |
| RVEDP, mmHg | 0.77 (0.27-2.22) | 0.630 | 1.45 (0.55-3.81) | 0.453 | **3.92 (1.27-12.09)** | **0.018** | 1.55 (0.87-2.76) | 0.138 |
| RVESP, mmHg | 2.16 (0.81-5.77) | 0.126 | **2.50 (1.02-6.14)** | **0.046** | **3.23 (1.24-8.42)** | **0.016** | **1.85 (1.09-3.13)** | **0.022** |
| P_max_, mmHg | 2.54 (0.87-7.41) | 0.089 | 2.34 (0.86-6.35) | 0.094 | **4.55 (1.32-15.71)** | **0.017** | **1.90 (1.08-3.36)** | **0.026** |
| dP/dt_max_, mmHg/s | 1.43 (0.52-3.93) | 0.490 | 2.63 (0.92-7.48) | 0.071 | **3.63 (1.08-12.18)** | **0.037** | 1.56 (0.90-2.69) | 0.110 |
| dP/dt_min_, mmHg/s | **0.21 (0.06-0.76)** | **0.017** | **0.27 (0.08-0.94)** | **0.040** | **0.07 (0.01-0.54)** | **0.010** | **0.29 (0.14-0.61)** | **0.001** |
| β | 0.83 (0.51-1.35) | 0.445 | 0.69 (0.44-1.07) | 0.580 | 0.88 (0.60-1.30) | 0.527 | 0.90 (0.73-1.12) | 0.344 |

##### Supplemental Table 4. Comparisons for Single-Beat Analysis Derived Hemodynamic Characteristics between Subgroups. Data are presented as mean ± SD. Hemodynamic and single beat analysis variables were compared between clusters using Mann-Whitney test.

| **Single Beat P-V Loop Variable** | **Group A (n=15)** | **Group B (n=24)** | **P-value** |
| --- | --- | --- | --- |
| Gold Standard | | | |
| Ees, mmHg/mL | 0.38 ± 0.13 | 0.34 ± 0.12 | 0.3408 |
| Ea, mmHg/mL | 1.16 ± 0.36 | 0.41 ± 0.20 | **<0.0001** |
| Ees/Ea, mmHg/mL | 0.38 ± 0.20 | 0.96 ± 0.45 | **<0.0001** |
| Eed, mmHg/mL | 0.045 ± 0.024 | 0.041 ± 0.025 | 0.7290 |
| Logistic Tau, ms | 32 ± 6 | 29 ± 4 | **0.0209** |
| RVEF % | 26 ± 10 | 47 ± 10 | **<0.0001** |
| RVEDV, mL | 286 ± 187 | 160 ± 53 | **0.0003** |
| RVESV, mL | 224 ± 184 | 86 ± 38 | **<0.0001** |
| RVEDP, mmHg | 13.3 ± 14.0 | 6.4 ± 3.7 | **0.0166** |
| RVESP, mmHg | 71 ± 23 | 27 ± 12 | **<0.0001** |
| P_max_, mmHg | 94 ± 22 | 50 ± 15 | **<0.0001** |
| dP/dt_max_, mmHg/s | 536 ± 225 | 365 ± 113 | **0.0014** |
| dP/dt_min_, mmHg/s | -726 ± 369 | -293 ± 109 | **<0.0001** |
| β | 0.037 ± 0.012 | 0.021 ± 0.014 | **0.0003** |
| Pipeline | | | |
| Ees, mmHg/mL | 0.35 ± 0.10 | 0.26 ± 0.11 | **0.0073** |
| Ea, mmHg/mL | 1.05 ± 0.27 | 0.41 ± 0.19 | **<0.0001** |
| Ees/Ea, mmHg/mL | 0.37 ± 0.15 | 0.76 ± 0.40 | **0.0002** |
| Eed, mmHg/mL | 0.035 ± 0.016 | 0.037 ± 0.024 | 0.7766 |
| Logistic Tau, ms | 39 ± 6 | 33 ± 4 | **0.0010** |
| RVEF % | 26 ± 8 | 41 ± 11 | **0.0002** |
| RVEDV, mL | 257 ± 74 | 185 ± 60 | **0.0067** |
| RVESV, mL | 195 ± 70 | 112 ± 46 | **0.0005** |
| RVEDP, mmHg | 8.5 ± 3.1 | 5.9 ± 3.4 | **0.0262** |
| RVESP, mmHg | 64 ± 13 | 27 ± 10 | **<0.0001** |
| P_max_, mmHg | 85 ± 13 | 44 ± 13 | **<0.0001** |
| dP/dt_max_, mmHg/s | 431 ± 65 | 245 ± 80 | **<0.0001** |
| dP/dt_min_, mmHg/s | -606 ± 116 | -285 ± 102 | **<0.0001** |
| β | 0.034 ± 0.007 | 0.022 ± 0.014 | **0.0020** |

##### Supplemental Table 5. Cardiac MRI measurements. Data are presented as mean ± SD.

| **Resting Cardiac MRI** | **Overall (n=39)** |
| --- | --- |
| LVEDV, mL | 129 ± 35 |
| LVESV, mL | 51 ± 17 |
| LVEDV/BSA, mL/m^2^ | 67 ± 14 |
| LVESV/BSA, mL/m^2^ | 26.2 ± 8.5 |
| LVEF, % | 60 ± 7 |
| LV Mass ED, g/m^2^ | 87 ± 23 |
| RVEDV, mL | 165 ± 55 |
| RVESV, mL | 83 ± 40 |
| RVEDV/BSA, mL/m^2^ | 87 ± 26 |
| RVESV/BSA, mL/m^2^ | 43 ± 20 |
| RVEF, % | 49 ± 10 |
| RV Mass ED, g/m^2^ | 32 ± 11 |

##### Supplemental Table 6. Goodness of Fit Metrics between Multi-Beat P-V Loop Variables and Pipeline Results. Data are presented as mean ± SD.

| **P-V Loop Variable** | **Mean** | **CCC** | **B** | **R** | **Mean Difference (± SD)** |
| --- | --- | --- | --- | --- | --- |
| Ees, mmHg/mL | 0.3 ± 0.1 | 0.189 | 0.38 | 0.95 | -0.38 ± 0.28 |
| Ea, mmHg/mL | 0.65 ± 0.38 | 0.871 | 0.92 | 0.97 | -0.03 ± 0.20 |
| Ees/Ea, mmHg/mL | 0.61 ± 0.38 | 0.346 | 0.50 | 0.88 | -0.54 ± 0.50 |
| RVEDV, mL | 213 ± 73 | 0.354 | 1.46 | 0.97 | 72 ± 55 |
| RVESV, mL | 144 ± 69 | 0.262 | 1.84 | 0.95 | 71 ± 57 |
| RVEDP, mmHg | 6.9 ± 3.5 | 0.001 | 0.01 | 0.21 | -39 ± 200 |
| dP/dt_max_, mmHg/s | 317 ± 118 | 0.287 | 0.64 | 0.96 | -178 ± 122 |
| dP/dt_min_, mmHg/s | -408 ± 191 | 0.777 | 0.81 | 0.98 | 95 ± 86 |

### SUPPLEMENTAL FIGURES

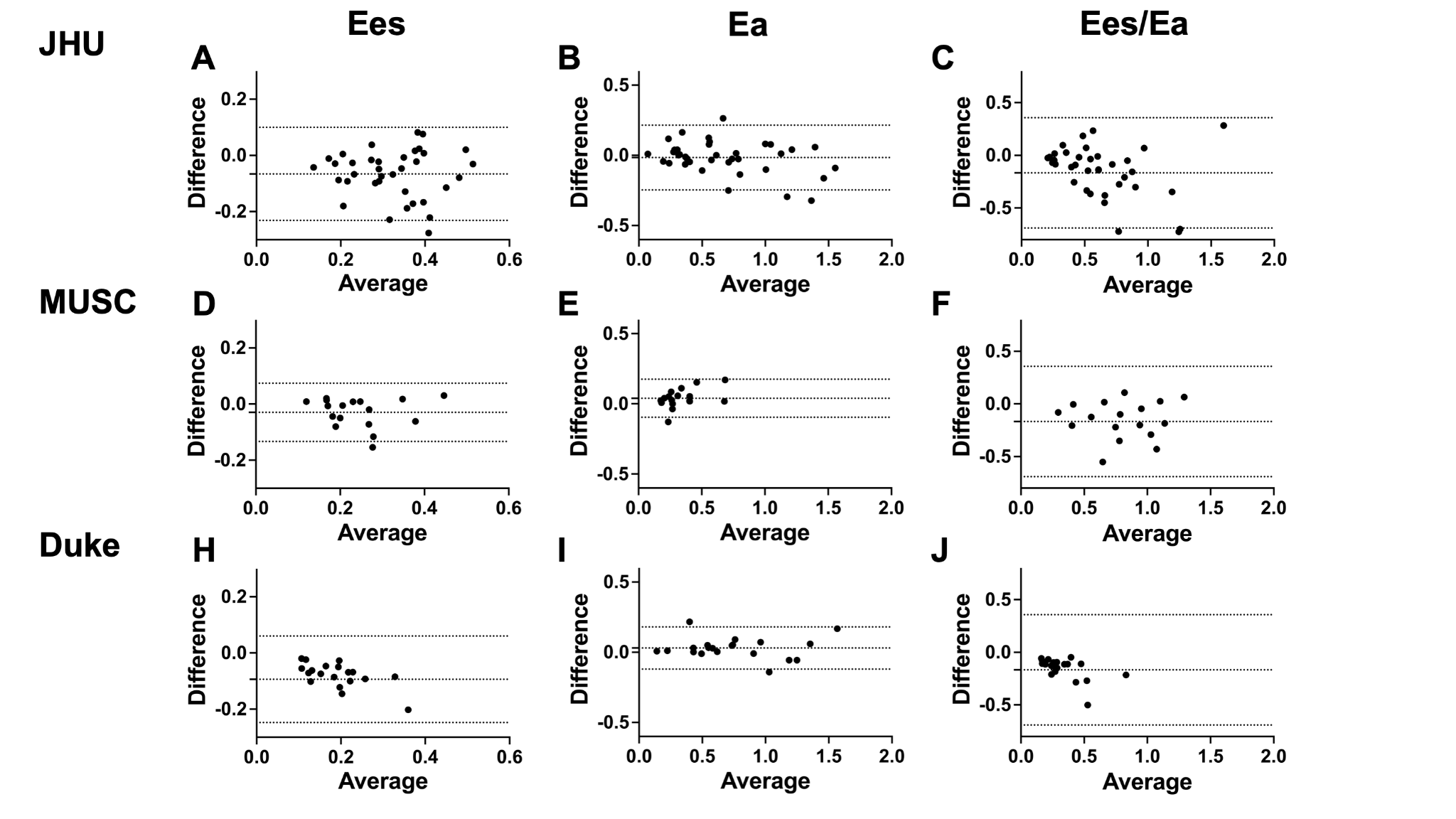

**Supplemental Figure 1. Bland-Altman analysis of Ea, Ees, Ees/Ea in different cohorts between gold standard and pipeline outputs.** Mean bias with 95% CIs is shown for all analyses.
